# Stable temporal relationships as a first step towards causal inference: an application to antibiotic resistance

**DOI:** 10.1101/2022.01.31.22270156

**Authors:** Avi Baraz, Michal Chowers, Daniel Nevo, Uri Obolski

**Affiliations:** School of Public Health, Tel Aviv University, Tel Aviv, Israel; Porter School of the Environment and Earth Sciences, Tel Aviv University, Tel Aviv, Israel; Department of Statistics and Operations Research, Tel Aviv University, Tel Aviv, Israel; Meir Medical Center, Kfar Saba, Israel; Sackler Faculty of Medicine, Tel Aviv University, Israel

## Abstract

Epidemiological studies often analyze data as static, essentially averaging observed associations across time. Overlooking time trends is especially problematic in settings subject to rapid changes. A prominent example for such a setting is antibiotic resistance, which has reached concerning levels, and poses a global healthcare challenge. Bacteria constantly evolve and hence antibiotic resistance is characterized by time-varying relationships with clinical and demographic covariates. In this paper, we speculate that covariates with a causal effect are expected to have stable relationships with resistance over calendar time. To this end, we applied time-varying coefficient models in a retrospective cohort analysis of a large clinical dataset from an Israeli hospital, and have shown their advantages in describing covariate-resistance relationships. We found both time-stable and time-varying covariate-resistance relationships. These results serve as initial evidence towards causal interpretation of these relationships, as one may expect time-stable rather than time-varying relationships to correspond with causal effects. We further conducted data-driven simulations, that have illustrated how results from time-varying coefficient models must be carefully interpreted with respect to causal claims. Potentially, identification of causal covariate-resistance relationships can lead to new medical interventions and healthcare policies, and improve the generalization of existing predictive models for antibiotic resistance.

## 1. Introduction

Antibiotic resistance is a major global healthcare concern. According to a 2019 report by the Centers for Disease Control and Prevention, in the US alone at least 2.8 million people suffer from antibiotic resistant bacterial infections each year, of which approximately 35,000 result in death (1). Antibiotic use and over-prescription have led to high global resistance levels, prompting the World Health Organization to declare antibiotic resistance a global health crisis, which also threatens the global economy (1, 2).

Antibiotic resistant infections emerge through complex biological processes, shaped by evolutionary forces and bacterial population dynamics (3, 4). The interplay between these processes and epidemiological-level covariates is still poorly understood. Moreover, estimates of epidemiological-level effects are likely to vary with time, due to the rapid dynamics of bacterial populations in response to environmental changes. For example, resistance frequencies are constantly changing as a result of selective pressure of antibiotic use in medicine (5–8) or agriculture (8–11), demographic changes such as urbanization (12, 13) and healthcare-related policies (14–16).

The causes of antibiotic resistance have been the focal point of much research. Randomized controlled trials, which can help unravel these causes, are in many cases difficult and impractical to perform in the context of antibiotic resistance. Hence, many of the studies that attempted to identify the causes of antibiotic resistance were observational. However, observational studies are subject to confounding, which limits their ability to determine if a risk factor causes antibiotic resistance or is only associated with it. Nonetheless, there have been some attempts, including by our group, to overcome confounding from observational studies to estimate the effect of antibiotic use on resistance (e.g. (17–21)).

In this work, we showcased that estimates of time-stable relationships (over calendar time) between clinical and demographic covariates and resistance, can provide evidence towards causality. If a covariate has a causal effect on an outcome variable, one may expect the relationship between them to be time-stable rather than time-varying. The estimated relationship estimate is not necessarily an estimate of the causal effect of the covariate on resistance, but rather evidence towards the existence of a causal relationship. We emphasize that this paper is concerned with calendar time-varying mechanisms resulting from the dynamic nature of infectious diseases. It is not concerned with time-varying treatments and confounders within an individual.

Demonstrating these claims empirically, we estimated temporal relationships between hospitalized patients’ bacterial antibiotic susceptibility test results and their corresponding electronic medical records during 2016-19. We modelled these temporal relationships by logistic time-varying coefficient models. Such models are a private case of Generalized Additive Models (GAM) (22, 23), that allows flexible modelling of non-linear covariate-outcome relationships in time. Previous research used GAMs in the context of antibiotic resistance. Two such papers dealt with the relationship between population-level antibiotic use and the emergence of resistance (24, 25), while another paper utilized GAMs, but not time-varying coefficient models, to model non-linear relationships between hospitalization duration and resistance (26).

We compared our results to standard time-fixed logistic regression models, and explored the differences between them. Our results presented significant time trends in the relationships between risk factors and resistance otherwise overlooked by standard models. Temporal relationships between risk factors and resistance that were estimated as stable, coincided with prior knowledge deeming them causal. On the other hand, temporal relationships estimated as time-varying corresponded to prior knowledge suggesting they were not causal. Finally, we conducted data-driven simulations that demonstrated the need for careful interpretation of obtained time-varying coefficients, as various plausible causal scenarios may give rise to observed time trends.

## 2. Methods

### 2.1 The motivating data

The dataset used in this retrospective cohort analysis includes electronic records of all patients who had a positive bacterial culture between 2016-19 in Meir Medical Center, Kfar Saba, Israel. The data originated from two types of records: Bacterial antibiotic susceptibility test results, and their corresponding patients’ demographic and clinical data.

Our analysis focused on resistance to gentamicin. Susceptibility testing to gentamicin was commonly performed, yielding a resistance rate of 15.6%, thus providing us a large sample with satisfactory event rate. Seven bacterial species commonly isolated in hospitalized patients *(Eescherichia coli, Pseudomonas aeruginosa, Klebsiella pneumoniae, Proteus mirabilis, Enterobacter cloacae, Morganella morganii and Citrobacter koseri)* were chosen for the analysis, since each had at least 200 resistance results to gentamicin. In total, 11,592 gentamicin resistance results from the above-mentioned bacterial species were sampled between 2016-19 and included in our analysis. To account for the heterogeneity of the species, a categorical covariate indicating the bacteria’s genus was incorporated in all our models. Table 1 presents summary statistics of key patient covariates, stratified by resistance test result (susceptible/resistant), and Tables S1 and S2 describe in detail and present summary statistics of all the covariates used in our analysis.

**Table 1.**
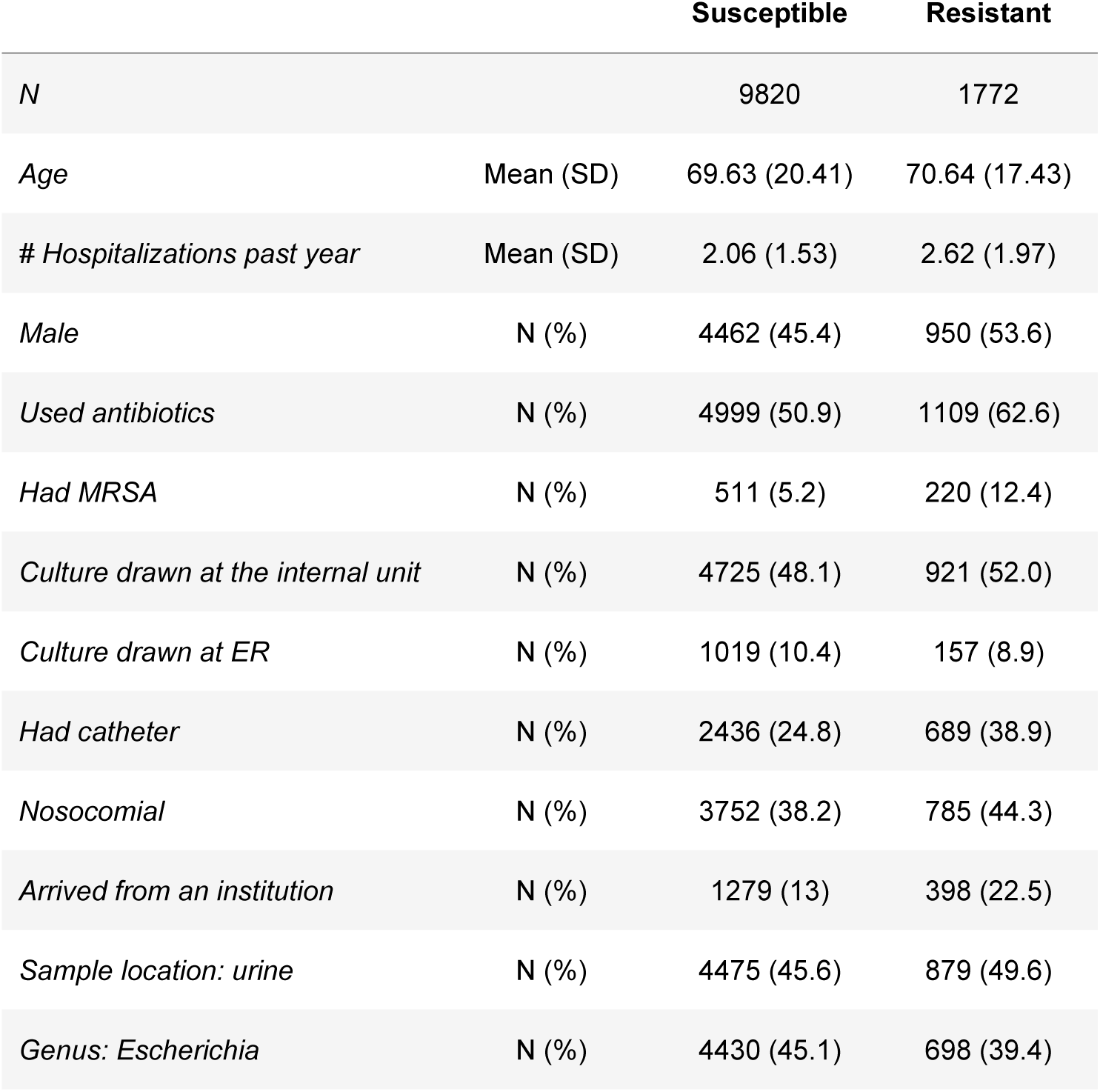
Summary of key covariates used in this study, stratified by gentamicin resistance result. #: number of. MRSA: Methicillin-resistant *Staphylococcus aureus*. ER: emergency room. *Used antibiotics*: hospital use of antibiotics inducing gentamicin resistance during the 6 months prior to the bacterial culture’s drawing.

|  |  | Susceptible | Resistant |
| --- | --- | --- | --- |
| *N* |  | 9820 | 1772 |
| *Age* | Mean (SD) | 69.63 (20.41) | 70.64 (17.43) |
| *# Hospitalizations past year* | Mean (SD) | 2.06 (1.53) | 2.62 (1.97) |
| *Male* | N (%) | 4462 (45.4) | 950 (53.6) |
| *Used antibiotics* | N (%) | 4999 (50.9) | 1109 (62.6) |
| *Had MRSA* | N (%) | 511 (5.2) | 220 (12.4) |
| *Culture drawn at the internal unit* | N (%) | 4725 (48.1) | 921 (52.0) |
| *Culture drawn at ER* | N (%) | 1019 (10.4) | 157 (8.9) |
| *Had catheter* | N (%) | 2436 (24.8) | 689 (38.9) |
| *Nosocomial* | N (%) | 3752 (38.2) | 785 (44.3) |
| *Arrived from an institution* | N (%) | 1279 (13) | 398 (22.5) |
| *Sample location: urine* | N (%) | 4475 (45.6) | 879 (49.6) |
| *Genus: Escherichia* | N (%) | 4430 (45.1) | 698 (39.4) |

### 2.2 Statistical methods

Temporal relationships between patients’ clinical and demographic covariates, and resistance, were modelled by logistic time-varying coefficient models, which are an extension of standard logistic regression models (22, 23). Unlike standard logistic regression models, time-varying coefficient models allow for variation of covariate-outcome relationships across time by replacing some of the time-fixed coefficients with non-parametric functions of time. These models take the form

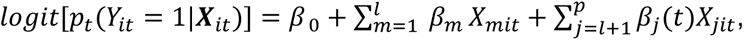

where *Y*_*it*_ is the gentamicin resistance, and ***X***_*it*_ = (*X*_1*it*_, . . ., *X*_*pit*_) is the vector of *p* covariates of the *i*^*th*^ observation, both measured at time *t*, which is the time when the bacterial culture was drawn from the patient (22, 23). We henceforth omit the index *t* from ***X***_*i*_ and *Y*_*i*_ for simplicity of presentation. The parameters of the model are the intercept *β*_0_, the time-fixed coefficients *β*_1_, . . ., *β*_*l*_, and the time-varying coefficients *β*_*l*+1_(*t*), . . ., *β*_*p*_(*t*). The model’s interpretation is that *e*^*β*(*t*)^ is the adjusted odds ratio (OR) between the resistance and a unit change in the *j* ^*th*^ variable at time *t*.

We modelled *β*_*l*+1_(*t*), . . ., *β*_*p*_(*t*) by penalized cubic splines with 40 equi-spaced knots, allowing them to be smooth functions of time. Smoothing parameters were estimated via Restricted Maximum Likelihood (REML), and subsequently *β*_0_ and ***β*** = (*β*_1_, . . ., *β*_*l*_, *β*_*l*+1_(*t*), . . ., *β*_*p*_(*t*)) ^*T*^ were estimated by minimizing the penalized model deviance (27).

Penalized splines can be expressed from a Bayesian perspective, which allows deriving credible intervals for them (27), while having frequentist coverage probabilities (28–30). Models including different sets of time-varying and time-fixed covariates were compared by a modified version of the Akaike Information Criterion (AIC) that accounts for the uncertainty in the smoothing parameter estimation (31, 32). The analysis was performed in R version 3.6.1 using the packages *mgcv* version 1.8-28 (29), *tableone* version 0.10.0 and *mgcViz* version 0.1.6.

The potential dependence between resistance results of bacteria from the same patient was addressed using random intercepts. The incorporation of random intercepts did not change the results substantially, and therefore the results of random-intercept models are presented in the supporting information.

### 2.3 Data-driven simulations

Estimated time-varying covariate-outcome relationships might misrepresent the true underlying causal effect. To further study this, two data-driven simulation studies were conducted. The simulations were designed with a confounding mechanism that affects coefficient estimates while following a hypothetical yet relevant clinical scenario.

In each simulation study, a Data Generating Mechanism (DGM) was designed to set up a scenario illustrating the challenges of interpreting estimated time-varying coefficients. The DGM determined hypothetical relationships between the 20 covariates previously included in our models and the outcome variable of gentamicin resistance. The simulations were data-driven in the sense that the values of the covariates in the DGM were taken from our data. Hence the simulated data structure, including the sample size, were comparable to our data. In each simulation scenario, all but one of the coefficients used to simulate the covariate-outcome relationships were the estimates from the models fitted to the original data. In addition, a hypothetical binary covariate denoted *Community use* was created and included in the DGM of both scenarios, and its values were sampled in each iteration of the simulation. We did not have access to data about antibiotic use outside the hospital. Therefore, we designated *Community use* to hypothetically indicate whether a patient was prescribed antibiotics outside the hospital during the year prior to their susceptibility test. The conditional probability of *Community use* was determined using Bayes’ theorem as detailed in the Results section.

Each simulation study consisted of 500 iterations. In each iteration, 11,592 gentamicin resistance results were drawn from the DGM. Then, a time-varying coefficient model with one time-varying coefficient (keeping the rest of the coefficients time-fixed (was fitted using the actual covariates in the data, the simulated values of Community use and resistance results. Finally, the average of the 500 time-varying coefficient estimates alongside their empirical 2.5% and 97.5% quantiles were calculated, as well as the average estimate and empirical 2.5% and 97.5% quantiles from an analogous standard (time-fixed) logistic regression model fitted to the same simulated data.

## 3. Results and discussion

### 3.1 Time-varying coefficient models applied to clinical data

Using our clinical data, we modelled the probability of resistance to gentamicin as a function of 20 covariates that were selected based on prior knowledge (33), and are detailed in Tables S1 and S2. We fitted one standard logistic regression model, and four logistic time-varying coefficient models, each with a different single time-varying coefficient.

The four coefficients allowed to vary in time correspond to different patient-level clinical and demographic information: whether the patient’s culture was drawn at the Emergency Room (ER) (*Culture drawn at ER*); the patient’s sex (*Male*); hospital use of relevant antibiotics during the prior 6 months (*Used antibiotics*), and whether the patient had a Methicillin-resistant *Staphylococcus aureus* (MRSA) infection during the prior year (*Had MRSA)*. Relevant antibiotics were considered those previously shown to have direct cross-resistance links with gentamicin (19). We correspondingly refer to these four models as the *Culture drawn at ER* model, *Male* model, *Used antibiotics* model and *Had MRSA* model. For example, in the *Culture drawn at ER* model, the coefficient of *Culture drawn at ER* was allowed to vary in time, as expressed by having *β*_1_(*t*) (and not *β*_1_) in the following model equation,

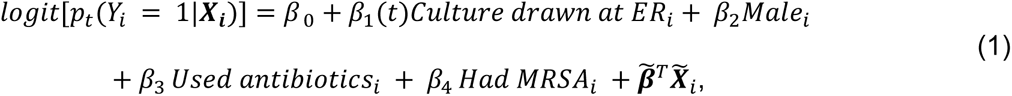

where *i* stands for the *i*^*th*^ observation, *Y*_*i*_ is the resistance, ***X***_*i*_ is the vector of all covariates, *β*_0_ is the intercept, *β*_1_(*t*), *β*_2_, *β*_3_, *β*_4_ are the coefficients of the listed covariates, and 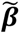 and 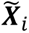 are the vectors of the rest of the 16 coefficients and covariates, respectively.

Table 2 and Figure 1 present estimated coefficients of selected covariates from the four time-varying models and from a standard logistic model using the same 20 covariates. The AIC values (Table 2) suggest that allowing the coefficients of *Culture drawn at ER* and *Male* to vary with time is preferable to keeping them constant. The estimates of these coefficients vary substantially with time (Figure 1), suggesting that perhaps both should be allowed to simultaneously vary with time. Indeed, a model that allowed the coefficients of both *Culture drawn at ER* and *Male* to vary with time had an AIC score of 8970.2, a score lower than the AIC scores of the other five models we fitted (Table 2). The estimates from this model were similar to the estimates from the *Culture drawn at ER* and *Male* models (data not shown). The estimated coefficients which were kept time-fixed in all models were consistent with prior evidence (33) (Tables S3-S7).

**Figure 1.**
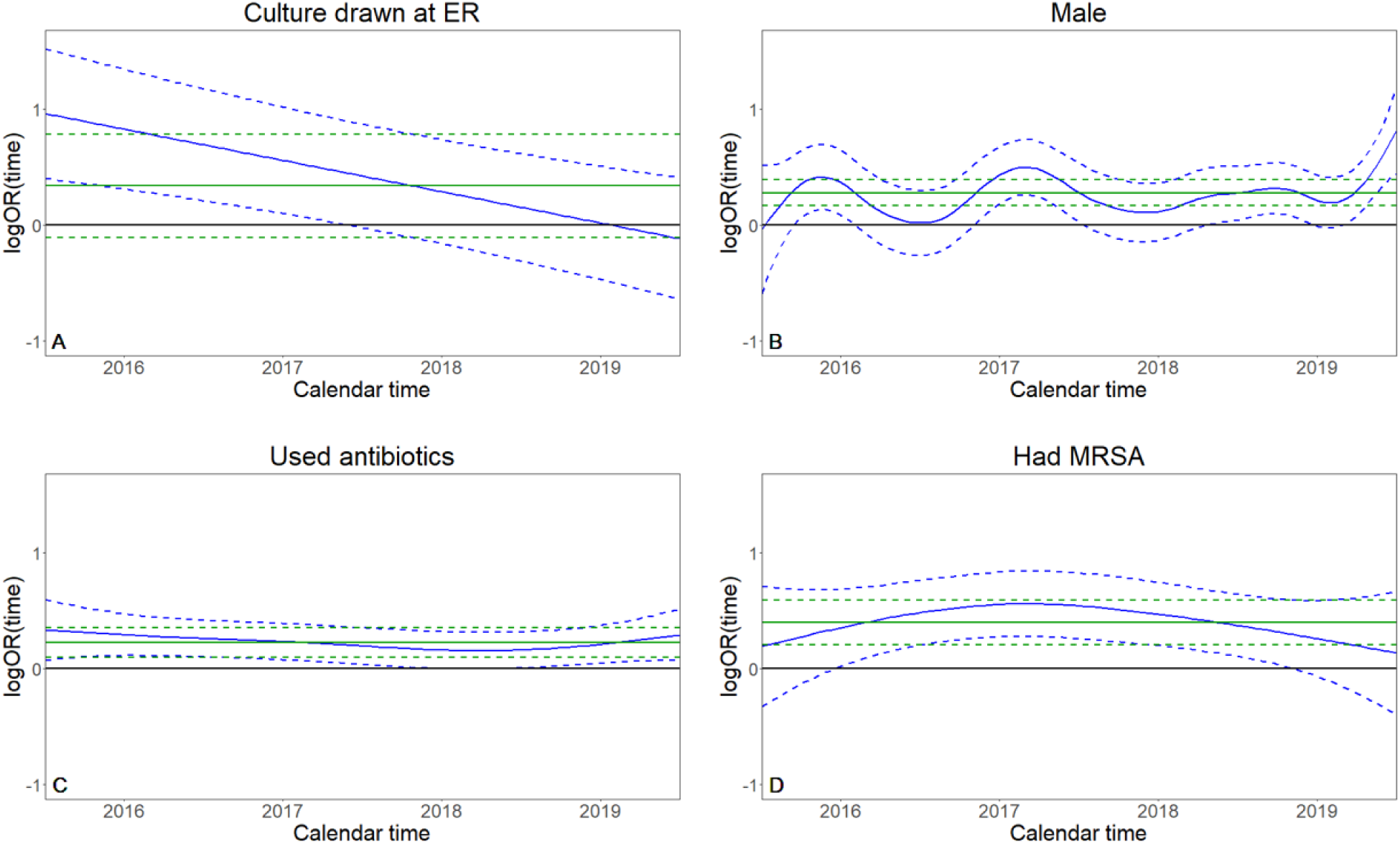
Estimates of the time-fixed (green) and time-varying (blue) coefficients from the single time-fixed and the four time-varying coefficient models. Correspondingly, 95% Bayesian credible intervals and standard 95% confidence intervals are given in dashed lines. The horizontal axis represents the time between 2016 and 2019.

**Table 2.** Selected coefficient estimates and AIC values for the five fitted models. In the standard logistic model, all coefficients were time-fixed. Each time-varying coefficient model was named after its time-varying coefficient, e.g. in the *Culture drawn at ER* model only the coefficient of *Culture drawn at ER* was time-varying. OR: odds ratio; CI: confidence interval; TV: the coefficient was time-varying; (/3): results presented per 3-unit increase.

|  | Standard<br>logistic model | *Culture drawn*<br>*at ER* model | *Male* model | *Used*<br>*antibiotics*<br>model | *Had MRSA*<br>model |
| --- | --- | --- | --- | --- | --- |
| Covariate | OR<br>(95% CI) |  |  |  |  |
| *Culture drawn at*<br>*ER* | 1.41<br>(0.91-2.19) | TV | 1.38<br>(0.88-2.13) | 1.41<br>(0.91-2.19) | 1.41<br>(0.91-2.19) |
| *Male* | 1.32<br>(1.18-1.48) | 1.32<br>(1.18-1.47) | TV | 1.32<br>(1.18-1.48) | 1.32<br>(1.18-1.48) |
| *Used antibiotics* | 1.26<br>(1.11-1.43) | 1.26<br>(1.11-1.44) | 1.25<br>(1.1-1.42) | TV | 1.26<br>(1.11-1.43) |
| *Had MRSA* | 1.49<br>(1.23-1.79) | 1.49<br>(1.23-1.80) | 1.51<br>(1.25-1.82) | 1.49<br>(1.23-1.80) | TV |
| *Times*<br>*hospitalized past*<br>*year (/3)* | 1.24<br>(1.11-1.38) | 1.24<br>(1.11-1.38) | 1.24<br>(1.11-1.39) | 1.25<br>(1.12-1.39) | 1.24<br>(1.11-1.38) |
| *Nosocomial* | 1.26<br>(1.12-1.43) | 1.27<br>(1.12-1.44) | 1.27<br>(1.12-1.44) | 1.26<br>(1.11-1.43) | 1.26<br>(1.11-1.43) |
| *Arrived from an*<br>*institution* | 1.43<br>(1.23-1.66) | 1.44<br>(1.24-1.67) | 1.43<br>(1.24-1.67) | 1.44<br>(1.24-1.67) | 1.43<br>(1.23-1.66) |
| (AIC) | (8990.1) | (8979.9) | (8981.0) | (8991.3) | (8991.3) |
| ΔAIC | 0 | -10.2 | -9.1 | +1.2 | +1.2 |

Considerable differences were observed between some of the estimated coefficients in the time-varying models and the standard logistic model (Figure 1A-B). While the standard logistic model yielded a positive coefficient estimate for *Culture drawn at ER*, the *Culture drawn at ER* model estimated it as positive only during 2016-18, with zero outside the credible interval only during 2016 to mid-2017, and with a decreasing linear trend towards the null/negative values with time (Figure 1A). Note that the obtained linear shape in Figure 1A was not a-priori imposed but resulted from the REML fitting procedure. The estimated *Male* coefficient also fluctuated with time in the *Male* model, but unlike the coefficient of *Culture drawn at ER,* no clear trend was observed (Figure 1B). When a random intercept was included in the *Male* model, a clearer mildly increasing trend was observed (Figure S1B). Of note is that both the time-varying and the standard logistic model estimated the coefficient of *Male* as positive during 2016-19.

On the other hand, the estimates of the time-varying coefficients in the *Used antibiotics* and *Had MRSA* models were similar to their corresponding estimates in the standard time-fixed logistic model, both in terms of point estimates and uncertainty bounds (confidence intervals for the time-fixed logistic model and credible intervals for the logistic time-varying coefficient models). With that being said, the estimated time-varying coefficient of *Had MRSA* slightly increased during 2016-2017 and then slightly decreased until the end of the study period.

The above-described results align with the arguments we pose in this work. Both *Used antibiotics* and *Had MRSA* are biologically plausible causes for antibiotic resistance. Prior antibiotic use causes resistance via evolutionary selective pressure (34), while a previous MRSA infection can imply remnants of resistant bacteria (35, 36). Thus, the estimated coefficients of *Used antibiotics* and *Had MRSA* demonstrate how time-stable relationships may align with causal interpretation. Conversely, it is biologically unlikely that *Culture drawn at ER* and *Male* are direct causes of resistant infections. Therefore, the estimated time-varying relationships of these covariates with resistance are more plausibly explained by confounding. We now turn to describe and illustrate such confounding mechanisms via data-driven simulations.

### 3.2 Exploring drivers of time-varying coefficient estimates via simulations

#### 3.2.1 First simulation study: Recreating a biased time-varying coefficient estimate

In the first simulation study, we recreated the time-varying coefficient estimate of *Culture drawn at ER* from the *Culture drawn at ER* model (Figure 1A), even though data were simulated such that *Culture drawn at ER* had no causal effect on resistance. That is, the underlying relationship was null. In this hypothetical scenario, the relationship between *Culture drawn at ER* and resistance was confounded by *Community use*, in a time-varying manner. In our scenario, we can hypothesize that increased awareness of antibiotic resistance led doctors treating outside the hospital to prescribe a decreasing number of antibiotics in the community during 2016-19, as was observed in various settings (37, 38). As a result, the association between *Culture drawn at ER* and *Community use* over time changed, so that *Culture drawn at ER* was a good surrogate for *Community use* at the beginning of 2016, but deteriorated over time.

The DGM for this scenario employed a standard time-fixed logistic model for the probability of resistance. The DGM included the same 20 covariates as the five models described in the previous subsection, apart from *Culture drawn at ER* which was replaced by *Community use*. For all covariates other than the artificially created *Community use*, the coefficients in the DGM were the estimates from the *Culture drawn at ER* model. We set the coefficient of *Community use* to two, as antibiotic use is a known cause for antibiotic resistance (34). In each simulation iteration, we simulated the resistance results according to a standard time-fixed logistic model

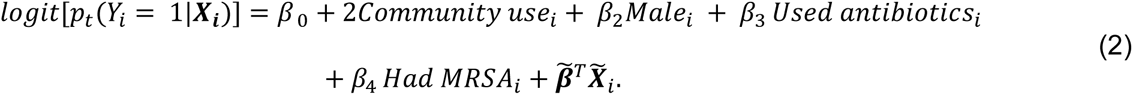

Using Bayes’ theorem, we set the probability of *Community use* as time-varying,

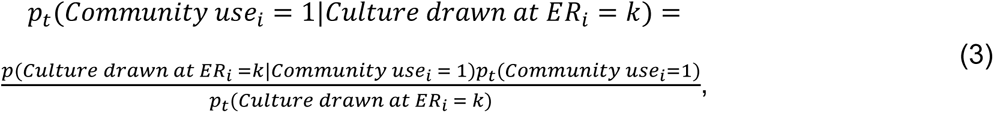

for *k* = 0,1. For simplicity, we chose a linear decrease of antibiotic prescription in the community over time, 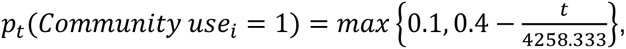 were chosen such that from t = 1279 (3.5 years) we obtained *p_t_*(*Community use_i_* = 1) = 0.1. The resulting approximate density of *p_t_*(*Community use_i_* = 1) in the data is shown in Figure S2. In our dataset, approximately 10% of the patients’ cultures were sampled at the ER. In this hypothetical scenario, patients who were prescribed antibiotics in the community were on average “less healthy”, and thus more likely to have arrived at the ER and had their bacterial culture drawn there, than patients who did not receive antibiotics in the community. Hence, we set *p_t_*(*Community drawn at ER_i_*|*Community use_i_* = 1) = 0.2. Finally, we estimated *p_t_*(*Community drawn at ER_i_* = 1) from the original data using a time-varying intercept-only model (Figure S3).

In each simulation iteration, we estimated a logistic time-varying coefficient model with *Culture drawn at ER* being the only time-varying coefficient (Methods). The average estimate of the time-varying coefficient of *Culture drawn at ER* across simulations (Figure 2) emulated the estimated coefficient obtained from the *Culture drawn at ER* model fitted to the original data (Figure 1A). To explain this, note that at the beginning of 2016 *Culture drawn at ER* was a good surrogate for *Community use*, and the average estimated time-varying coefficient was close to the actual coefficient of *Community use* in Equation (2) (Figure 2). Then, as the association between the two covariates diminished over time due to the trend of decrease in *Community use*, the average time-varying coefficient estimate of *Culture drawn at ER* diverged from the coefficient of *Community use* towards zero, its true value under the DGM (Equation (2)). Thus, this simulation study demonstrated how a confounding mechanism that varies with time can result in a misleading estimate.

**Figure 2.**
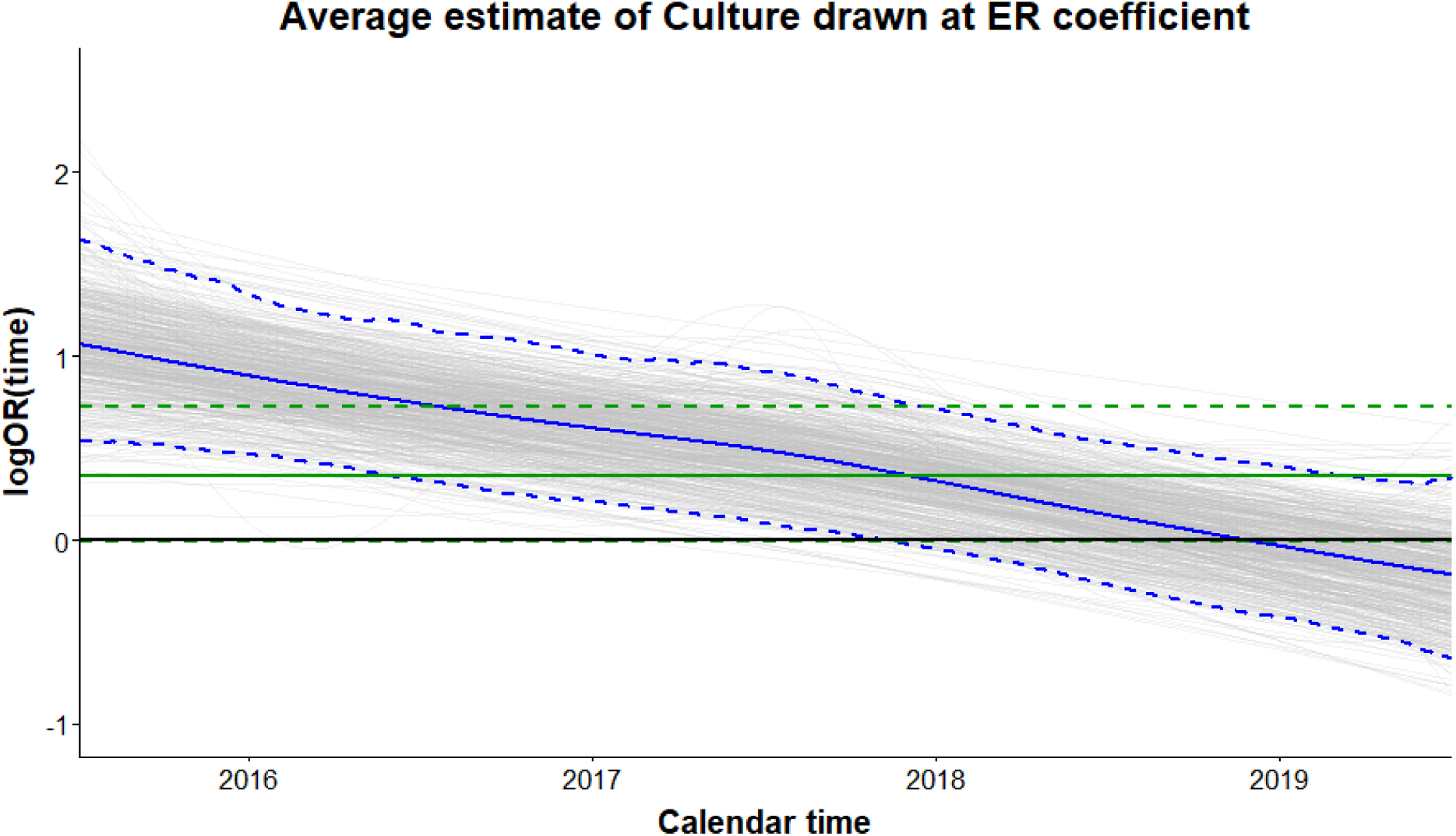
Average estimates of the time-fixed (green) and time-varying (blue) coefficients from the 500 time-fixed and time-varying coefficient models from the first simulation study. Correspondingly, 2.5% and 97.5% quantiles of coefficients estimates are in dashed green and blue. The 500 time-varying coefficient estimates are depicted in grey. The horizontal axis represents the time between 2016 and 2019.

For comparison, the average coefficient estimate of *Culture drawn at ER* from the standard time-fixed logistic model (Figure 2) was also biased. Its average was 0.35 (2.5%, 97.5% quantiles: - 0.01, 0.72), while in practice *Culture drawn at ER* had no effect on the resistance. Hence time-fixed models are also subject to bias in such a scenario.

#### 3.2.2 Second simulation study: Recreating a biased time-stable coefficient estimate

In the second data-driven simulation, we considered a causal relationship that varied considerably with time but was approximately estimated as time-stable, even though the estimated coefficient was allowed to vary in time. To obtain this approximate time-stable estimate, the confounding mechanism had to vary with time in a way that near-perfectly negated the underlying time-varying relationship. Hence, it demanded parameter values that are unlikely to be realistic in our context.

In this simulation scenario, the relationship between *Had MRSA* and resistance was confounded by *Community use* in a time-varying manner. The time-varying effect of *Had MRSA* on resistance was set as positive, and decreased over time. The decrease over time can be motivated through changes in the bacterial population of *Staphylococcus aureus*, as it can affect the resistance of other bacteria via horizontal gene transfer (HGT) (35). As in the first simulation, we mimicked increased awareness of antibiotic resistance that led community doctors to decrease antibiotic prescription over time. As a result, the association between *Had MRSA* and *Community use* changed over time, so that *Had MRSA* started as a good surrogate for *Community use* but deteriorated over time. We designed the change in the association between *Had MRSA* and *Community use* to specifically induce bias into the estimate of the time-varying coefficient of *Had MRSA*, such that a constant estimate was obtained.

We set the coefficient of *Community use* to 1.25, and the time-varying coefficient of *Had MRSA* to 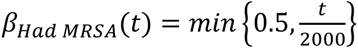. Then, in each iteration we simulated the resistance results according to the following DGM,

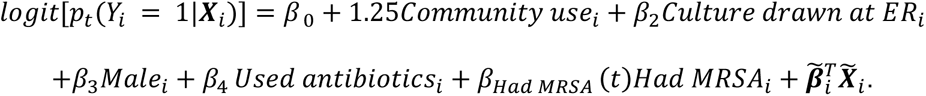

Again, we set the probability of *Community use* using Bayes’ Theorem,

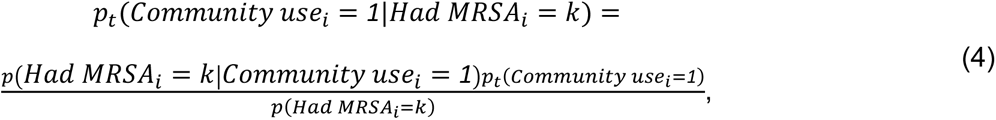

for *k* = 0,1. As in the previous simulation study, the values of each component in Equation (4) followed a clinical scenario in which antibiotic community use linearly decreased during 2016-19. Therefore, we set 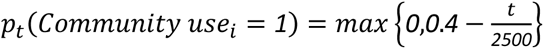 . In our dataset, approximately 6% of the patients have previously had an MRSA infection, hence we set *p*(*Had MRSA_i_* = 1) = 0.06. Finally, we assumed that patients who were prescribed antibiotics in the community are on average more likely to have had an MRSA infection than those who were not, and set *p*(*Had MRSA_i_*|*Community use_i_* = *1*) = *0*.*12*.

In each iteration, we estimated a logistic time-varying coefficient model with the 20 available covariates in which the coefficient of *Had MRSA* was the only time-varying coefficient (Methods section). The obtained average coefficient estimate (Figure 3) was time-stable, and resembled the coefficient estimates from the *Used antibiotics* and *Had MRSA* models in the original data (Figure 1C-D). As in the first simulation study, in each iteration we also fitted a standard time-fixed logistic model. The average estimate of the time-fixed coefficient of *Had MRSA* was also biased (Figure 3).

**Figure 3.**
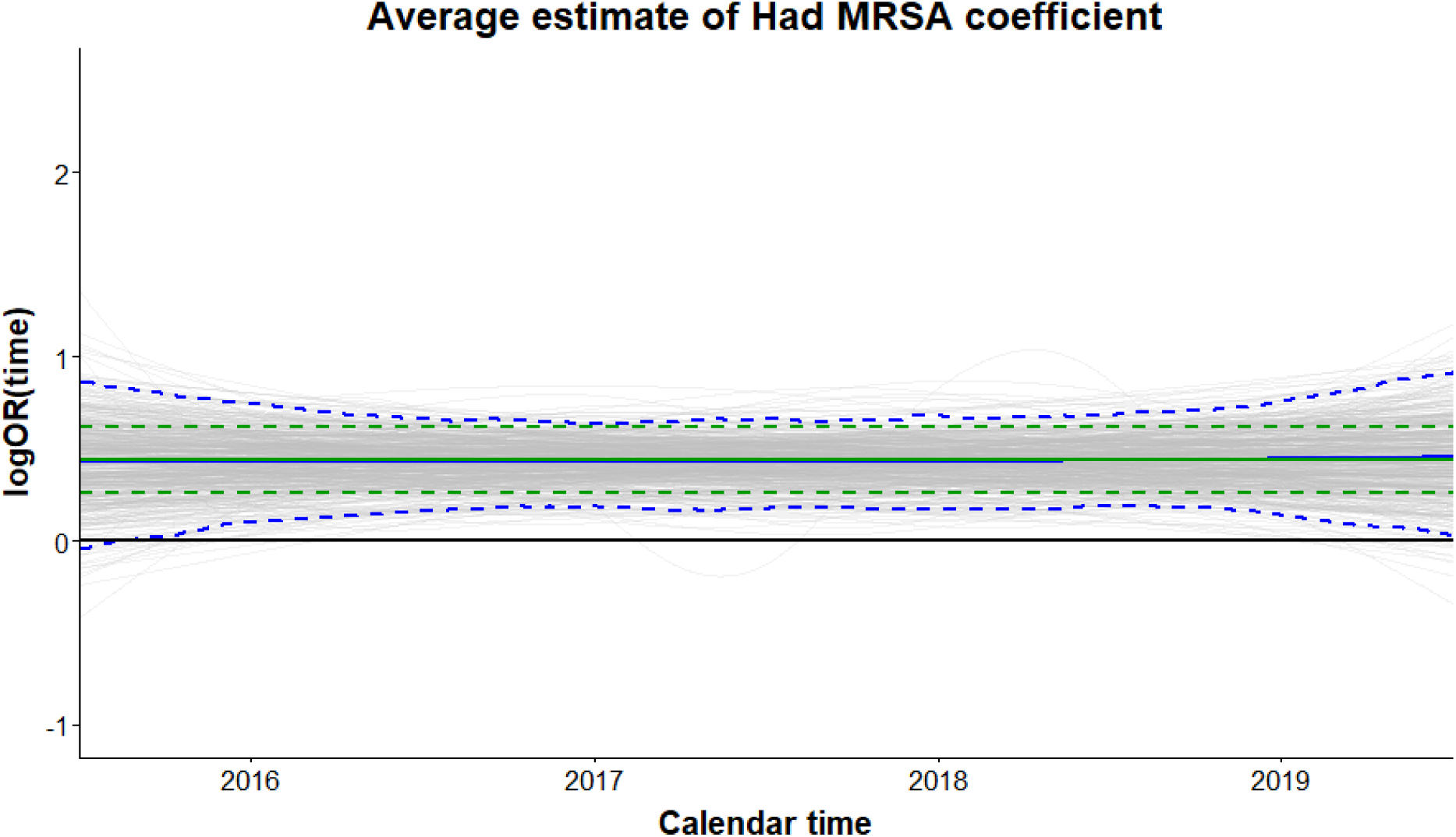
Average estimates of the time-fixed (green) and time-varying (blue) coefficients from the 500 time-fixed and time-varying coefficient models. Correspondingly, 2.5% and 97.5% quantiles of coefficients estimates are in dashed green and blue. The 500 time-varying coefficient estimates are depicted in grey. The horizontal axis represents the time between 2016 and 2019.

## Conclusions

Many epidemiological settings, and especially those involving infectious diseases, are characterized by time-varying relationships. However, epidemiological studies often analyze data as static, essentially averaging observed associations across time and neglecting to account for the dynamical nature of the studied system. Using a combination of an antibiotic resistance related clinical dataset and data-driven simulations, we have demonstrated that time-varying coefficient models can reveal phenomena otherwise obscured by time-fixed models.

We showcased how the use of time-varying coefficient models may help unravel causal covariate-outcome relationships. As we have shown, estimates that are time-stable can coincide with prior knowledge suggesting that the underlying relationships are causal. Conversely, time-varying estimates may direct researchers to inquire the underlying causal structure leading to the obtained time-varying relationships.

Time-varying coefficient estimates can result from a mixture of underlying covariate-outcome relationships and confounding mechanisms that are time-fixed and/or time-varying. It is possible the underlying covariate-outcome relationships are time-fixed, while time-varying confounding leads to a time-varying estimate; we demonstrated such a scenario in our first data-driven simulation study. On the other hand, time-stable coefficient estimates are more likely the consequence of underlying time-fixed covariate-outcome relationships, that are possibly confounded in a time-fixed manner. While it is technically possible that time-varying confounding will lead to a time-stable estimate of an underlying time-varying covariate-outcome relationship, this scenario is unlikely. Such a scenario requires the variations in the underlying relationship and in the confounding mechanism to approximately negate each other, as we illustrated in the second data-driven simulation study. This is implausible and hence can often be ruled out, though it is advised to do so based on subject-matter expertise.

From a clinical standpoint, knowledge about causal relationships can help researchers understand the biological mechanisms behind them, and possibly result in new interventions. This knowledge can also be exploited to improve existing predictive models for antibiotic resistance (e.g. (39–42)). By emphasizing covariates with causal effects on resistance, predictions from such models might prove more stable and robust to generalizations over different time periods or locations.

In the context of antibiotic resistance, future research should extend efforts of further identifying causal relationships between risk factors and antibiotic resistance using time-varying models. Development of a theoretical framework for assessing causal effects in the field of antibiotic resistance (i.e., based on potential outcomes and/or directed acyclic graphs), while accounting for variations with time, would be clinically and methodologically valuable.

## Data Availability

Data produced in the present study are proprietary but can be available upon reasonable request to the authors.

---

**Table S1.** Summary of the covariates used in this study, stratified by gentamicin resistance results. #: number of. MRSA: Methicillin-resistant *Staphylococcus aureus*. ER: emergency room. *Used antibiotics*: hospital use of antibiotics inducing gentamicin resistance during the 6 months prior to the bacterial culture’s drawing.

|  |  | Susceptible | Resistant |
| --- | --- | --- | --- |
| *N* |  | 9820 | 1772 |
| *Age* | Mean (SD) | 69.63 (20.41) | 70.64 (17.43) |
| *# Hospitalizations past year* | Mean (SD) | 2.06 (1.53) | 2.62 (1.97) |
| *Male* | N (%) | 4462 (45.4) | 950 (53.6) |
| *Used antibiotics* | N (%) | 4999 (50.9) | 1109 (62.6) |
| *Had MRSA* | N (%) | 511 (5.2) | 220 (12.4) |
| *Had Acinetobacter* | N (%) | 66 (0.7) | 31 (1.7) |
| *Had Pseudomonas* | N (%) | 116 (1.2) | 62 (3.5) |
| *Had CRE* | N (%) | 34 (0.3) | 16 (0.9) |
| *Had VRE* | N (%) | 19 (0.2) | 9 (0.5) |
| *Had ESBL* | N (%) | 1551 (15.8) | 697 (39.3) |
| *Had KPC* | N (%) | 109 (1.1) | 50 (2.8) |
| *Culture drawn at the internal unit* | N (%) | 4725 (48.1) | 921 (52.0) |
| *Culture drawn at the surgical unit* | N (%) | 2374 (24.2) | 386 (21.8) |
| *Culture drawn at the orthopedic unit* | N (%) | 983 (10) | 227 (12.8) |
| *Culture drawn at the obstetrics unit* | N (%) | 188 (1.9) | 22 (1.2) |
| *Culture drawn at another unit* | N (%) | 531 (5.4) | 59 (3.3) |
| *Culture drawn at ER* | N (%) | 1019 (10.4) | 157 (8.9) |
| *Had catheter* | N (%) | 2436 (24.8) | 689 (38.9) |
| *Nosocomial* | N (%) | 3752 (38.2) | 785 (44.3) |
| *Arrived from home* | N (%) | 5997 (61.1) | 942 (53.2) |
| *Arrived from an institution* | N (%) | 1279 (13) | 398 (22.5) |
| *Arrived from a medical clinic* | N (%) | 70 (0.7) | 14 (0.8) |
| *Arrived from another hospital* | N (%) | 256 (2.6) | 72 (4.1) |
| *Arrived from: unknown* | N (%) | 1511 (15.4) | 216 (12.2) |
| *Arrived from: other* | N (%) | 707 (7.2) | 130 (7.3) |
| *Diabetes* | N (%) | 1599 (16.3) | 354 (20) |
| *Immunosuppression* | N (%) | 307 (4) | 84 (4.7) |
| *Sample location: urine* | N (%) | 4475 (45.6) | 879 (49.6) |
| *Sample location: sputum* | N (%) | 706 (7.2) | 85 (4.8) |
| *Sample location: wound* | N (%) | 2045 (20.8) | 443 (25) |
| *Sample location: blood* | N (%) | 1126 (11.5) | 182 (10.3) |
| *Sample location: other* | N (%) | 63 (0.6) | 5 (0.3) |
| *Sample location: unknown* | N (%) | 1405 (14.3) | 178 (10) |
| *Genus: Escherichia* | N (%) | 4430 (45.1) | 698 (39.4) |
| *Genus: Entrobacter* | N (%) | 443 (4.5) | 31 (1.7) |
| *Genus: Citrobacter* | N (%) | 326 (3.3) | 5 (0.3) |
| *Genus: Pseudomonas* | N (%) | 2050 (20.9) | 304 (17.2) |
| *Genus: Proteus* | N (%) | 848 (8.6) | 273 (15.4) |
| *Genus: Klebsiella* | N (%) | 1411 (14.4) | 410 (23.1) |
| *Genus: Morganella* | N (%) | 312 (3.2) | 51 (2.9) |

**Table S2.**
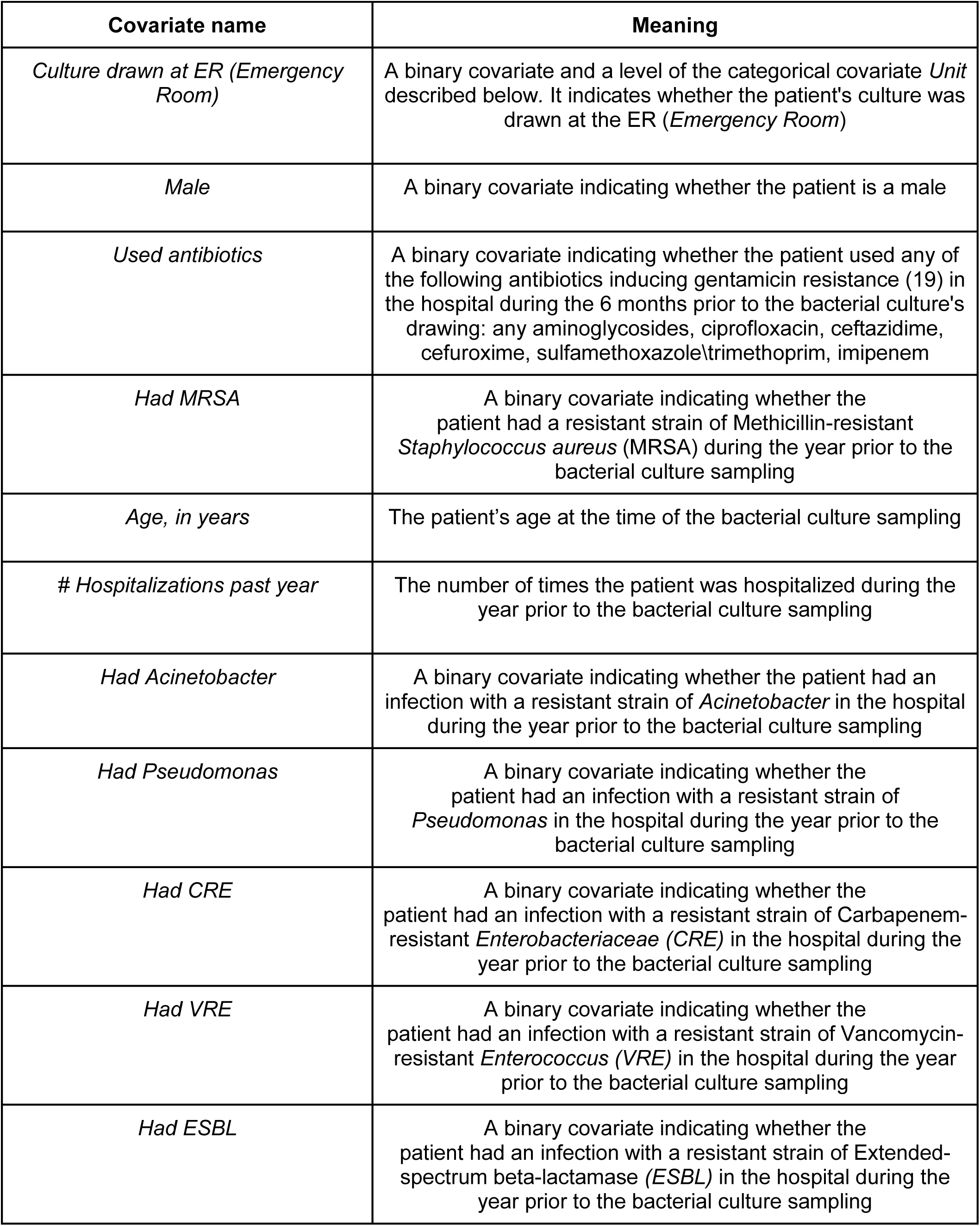
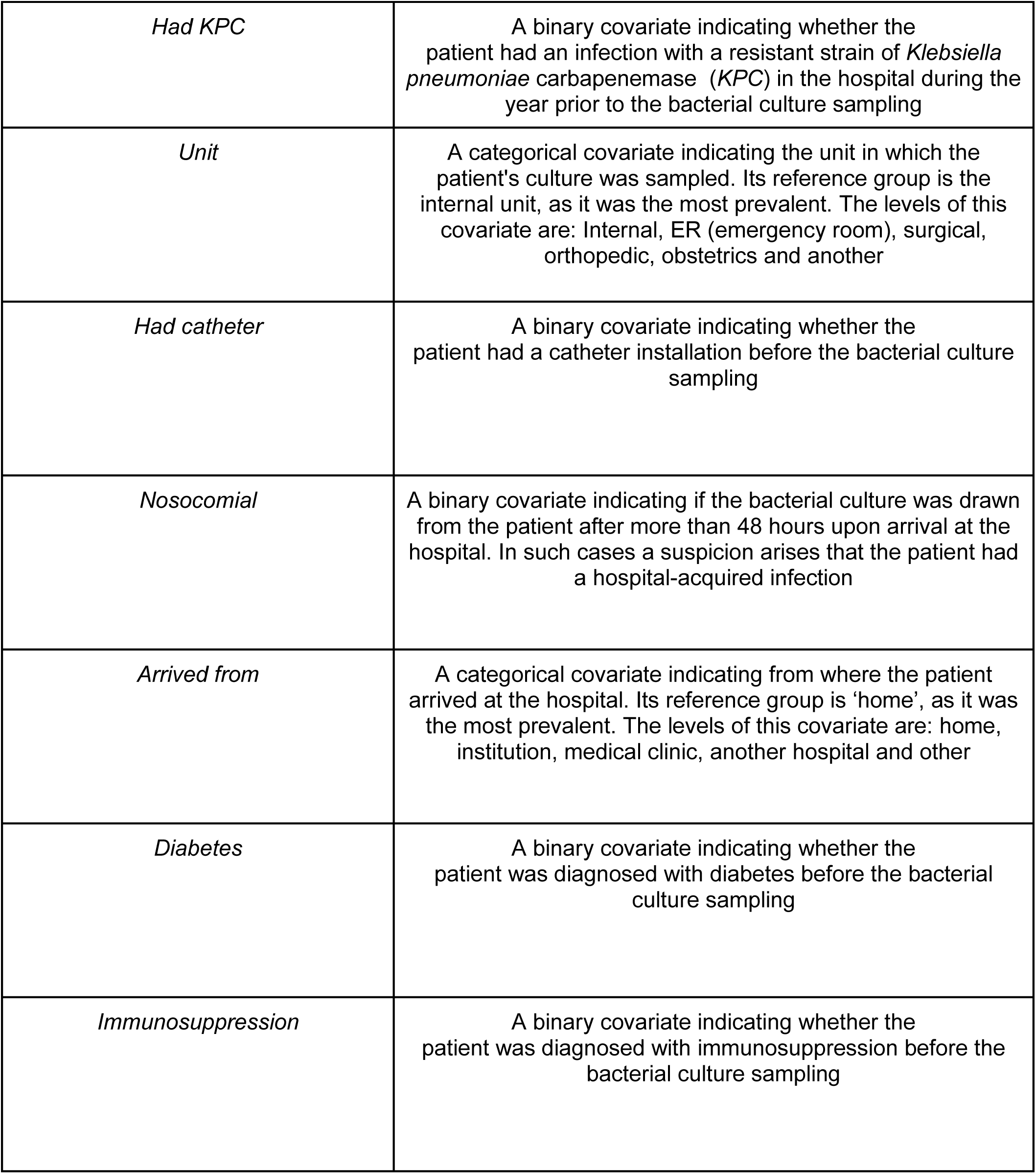
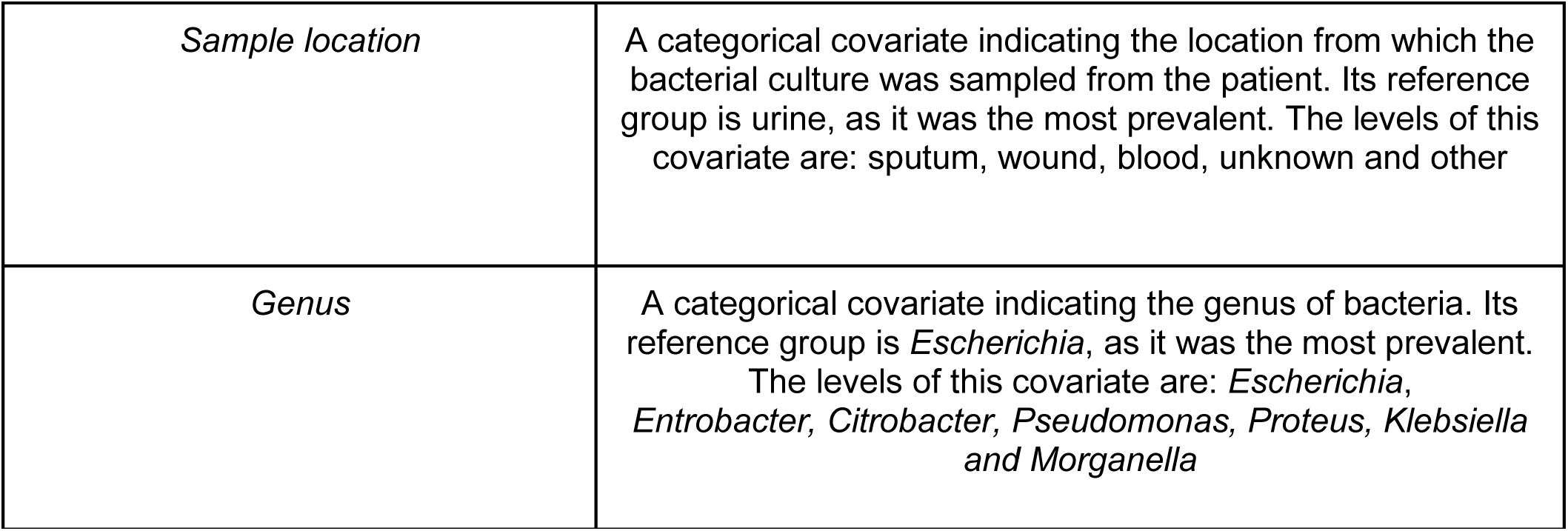
Description of the covariates used in this study. #: number of.

| Covariate name | Meaning |
| --- | --- |
| *Culture drawn at ER (Emergency Room)* | A binary covariate and a level of the categorical covariate *Unit* described below. It indicates whether the patient's culture was drawn at the ER (*Emergency Room*) |
| *Male* | A binary covariate indicating whether the patient is a male |
| *Used antibiotics* | A binary covariate indicating whether the patient used any of the following antibiotics inducing gentamicin resistance (19) in the hospital during the 6 months prior to the bacterial culture's drawing: any aminoglycosides, ciprofloxacin, ceftazidime, cefuroxime, sulfamethoxazole\trimethoprim, imipenem |
| *Had MRSA* | A binary covariate indicating whether the patient had a resistant strain of Methicillin-resistant *Staphylococcus aureus* (MRSA) during the year prior to the bacterial culture sampling |
| *Age, in years* | The patient's age at the time of the bacterial culture sampling |
| *# Hospitalizations past year* | The number of times the patient was hospitalized during the year prior to the bacterial culture sampling |
| *Had Acinetobacter* | A binary covariate indicating whether the patient had an infection with a resistant strain of *Acinetobacter* in the hospital during the year prior to the bacterial culture sampling |
| *Had Pseudomonas* | A binary covariate indicating whether the patient had an infection with a resistant strain of *Pseudomonas* in the hospital during the year prior to the bacterial culture sampling |
| *Had CRE* | A binary covariate indicating whether the patient had an infection with a resistant strain of Carbapenem-resistant *Enterobacteriaceae* (CRE) in the hospital during the year prior to the bacterial culture sampling |
| *Had VRE* | A binary covariate indicating whether the patient had an infection with a resistant strain of Vancomycin-resistant *Enterococcus* (VRE) in the hospital during the year prior to the bacterial culture sampling |
| *Had ESBL* | A binary covariate indicating whether the patient had an infection with a resistant strain of Extended-spectrum beta-lactamase (ESBL) in the hospital during the year prior to the bacterial culture sampling |
| *Had KPC* | A binary covariate indicating whether the patient had an infection with a resistant strain of *Klebsiella pneumoniae* carbapenemase (*KPC*) in the hospital during the year prior to the bacterial culture sampling |
| *Unit* | A categorical covariate indicating the unit in which the patient's culture was sampled. Its reference group is the internal unit, as it was the most prevalent. The levels of this covariate are: Internal, ER (emergency room), surgical, orthopedic, obstetrics and another |
| *Had catheter* | A binary covariate indicating whether the patient had a catheter installation before the bacterial culture sampling |
| *Nosocomial* | A binary covariate indicating if the bacterial culture was drawn from the patient after more than 48 hours upon arrival at the hospital. In such cases a suspicion arises that the patient had a hospital-acquired infection |
| *Arrived from* | A categorical covariate indicating from where the patient arrived at the hospital. Its reference group is 'home', as it was the most prevalent. The levels of this covariate are: home, institution, medical clinic, another hospital and other |
| *Diabetes* | A binary covariate indicating whether the patient was diagnosed with diabetes before the bacterial culture sampling |
| *Immunosuppression* | A binary covariate indicating whether the patient was diagnosed with immunosuppression before the bacterial culture sampling |
| *Sample location* | A categorical covariate indicating the location from which the bacterial culture was sampled from the patient. Its reference group is urine, as it was the most prevalent. The levels of this covariate are: sputum, wound, blood, unknown and other |
| *Genus* | A categorical covariate indicating the genus of bacteria. Its reference group is *Escherichia*, as it was the most prevalent. The levels of this covariate are: *Escherichia*, *Enterobacter*, *Citrobacter*, *Pseudomonas*, *Proteus*, *Klebsiella* and *Morganella* |

**Table S3.** Estimates of the standard logistic model fitted to the original data. (/3): results presented per 3 unit increase in the covariate; (/5): results presented per 5 unit increase in the covariate.

| *Covariate* | **Standard logistic model** |  |  |
| --- | --- | --- | --- |
|  | *Odds Ratios* | *Conf. Int (95%)* | *P-Value* |
| (Intercept) | 0.09 | 0.07 – 0.12 | <0.001 |
| Age (/5) | 0.98 | 0.97 – 1.00 | 0.028 |
| Times hospitalized past year (/3) | 1.24 | 1.11 – 1.38 | <0.001 |
| Had Acinetobacter | 2.05 | 1.23 – 3.35 | 0.005 |
| Had Pseudomonas | 1.95 | 1.38 – 2.74 | <0.001 |
| Had CRE | 1.54 | 0.78 – 2.90 | 0.194 |
| Had VRE | 0.64 | 0.25 – 1.54 | 0.335 |
| Had ESBL | 2.63 | 2.33 – 2.96 | <0.001 |
| Had KPC | 1.80 | 1.23 – 2.60 | 0.002 |
| Culture drawn at the surgical unit | 0.95 | 0.82 – 1.12 | 0.562 |
| Culture drawn at the orthopedic unit | 1.40 | 1.13 – 1.74 | 0.002 |
| Culture drawn at the obstetrics unit | 1.13 | 0.61 – 2.07 | 0.692 |
| Culture drawn at other unit | 0.65 | 0.46 – 0.90 | 0.011 |
| Had catheter | 1.26 | 1.09 – 1.44 | 0.001 |
| Nosocomial | 1.26 | 1.12 – 1.43 | <0.001 |
| Arrived from an institution | 1.43 | 1.23 – 1.66 | <0.001 |
| Arrived from a medical clinic | 0.96 | 0.50 – 1.73 | 0.885 |
| Arrived from another hospital | 1.33 | 0.97 – 1.80 | 0.069 |
| Arrived from: unknown | 0.99 | 0.66 – 1.46 | 0.943 |
| Arrived from: other | 0.97 | 0.78 – 1.20 | 0.767 |
| Diabetes | 1.02 | 0.89 – 1.18 | 0.775 |
| Immunosuppression | 1.23 | 0.95 – 1.58 | 0.115 |
| Sample location: unknown | 0.64 | 0.53 – 0.78 | <0.001 |
| Sample location: sputum | 0.45 | 0.34 – 0.59 | <0.001 |
| Sample location: wound | 0.96 | 0.81 – 1.14 | 0.670 |
| Sample location: blood | 0.78 | 0.65 – 0.93 | 0.007 |
| Sample location: other | 0.72 | 0.24 – 1.69 | 0.493 |
| Genus: Entrobacter | 0.46 | 0.30 – 0.66 | <0.001 |
| Genus: Citrobacter | 0.10 | 0.04 – 0.22 | <0.001 |
| Genus: Pseudomonas | 0.81 | 0.69 – 0.95 | 0.011 |
| Genus: Proteus | 1.60 | 1.35 – 1.90 | <0.001 |
| Genus: Klebsiella | 1.62 | 1.40 – 1.86 | <0.001 |
| Genus: Morganella | 0.80 | 0.57 – 1.11 | 0.194 |
| Used antibiotics | 1.26 | 1.11 – 1.43 | <0.001 |
| Had MRSA | 1.49 | 1.23 – 1.79 | <0.001 |
| Male | 1.32 | 1.18 – 1.48 | <0.001 |
| Culture drawn at ER | 1.41 | 0.91 – 2.19 | 0.126 |
| Observations | 11592 |  |  |

**Table S4.** Estimates of *Culture drawn at ER* model fitted to the orginal data. S(Culture drawn at ER): The time-varying coefficient of *Culture drawn at ER,* testing if this coefficient equals zero for all *t;* (/3): results presented per 3 unit increase in the covariate; (/5): results presented per 5 unit increase in the covariate.

| *Covariate* | **Culture drawn at ER model** |  |  |
| --- | --- | --- | --- |
|  | *Odds Ratios* | *Conf. Int (95%)* | *P-Value* |
| Intercept | 0.09 | 0.07 – 0.12 | <0.001 |
| Age (/5) | 0.98 | 0.97 – 1.00 | 0.017 |
| Times hospitalized past year (/3) | 1.24 | 1.11 – 1.38 | <0.001 |
| Had Acinetobacter | 2.07 | 1.25 – 3.40 | 0.004 |
| Had Pseudomonas | 1.96 | 1.39 – 2.77 | <0.001 |
| Had CRE | 1.54 | 0.80 – 2.96 | 0.191 |
| Had VRE | 0.64 | 0.26 – 1.57 | 0.329 |
| Had ESBL | 2.61 | 2.32 – 2.95 | <0.001 |
| Had KPC | 1.79 | 1.23 – 2.61 | 0.002 |
| Culture drawn at the surgical unit | 0.97 | 0.83 – 1.13 | 0.672 |
| Culture drawn at the orthopedic unit | 1.41 | 1.14 – 1.76 | 0.002 |
| Culture drawn at the obstetrics unit | 1.13 | 0.61 – 2.08 | 0.697 |
| Culture drawn at other unit | 0.65 | 0.46 – 0.90 | 0.010 |
| Had catheter | 1.26 | 1.10 – 1.45 | 0.001 |
| Nosocomial | 1.27 | 1.12 – 1.44 | <0.001 |
| Arrived from an institution | 1.44 | 1.24 – 1.67 | <0.001 |
| Arrived from a medical clinic | 0.95 | 0.51 – 1.77 | 0.875 |
| Arrived from another hospital | 1.33 | 0.98 – 1.82 | 0.067 |
| Arrived from: unknown | 0.99 | 0.66 – 1.47 | 0.955 |
| Arrived from: other | 0.97 | 0.78 – 1.20 | 0.767 |
| Diabetes | 1.03 | 0.90 – 1.19 | 0.656 |
| Immunosuppression | 1.23 | 0.95 – 1.59 | 0.114 |
| Sample location: unknown | 0.61 | 0.50 – 0.74 | <0.001 |
| Sample location: sputum | 0.45 | 0.34 – 0.59 | <0.001 |
| Sample location: wound | 0.95 | 0.80 – 1.12 | 0.539 |
| Sample location: blood | 0.77 | 0.64 – 0.93 | 0.005 |
| Sample location: other | 0.71 | 0.27 – 1.85 | 0.487 |
| Genus: Entrobacter | 0.46 | 0.31 – 0.67 | <0.001 |
| Genus: Citrobacter | 0.10 | 0.04 – 0.25 | <0.001 |
| Genus: Pseudomonas | 0.80 | 0.68 – 0.95 | 0.009 |
| Genus: Proteus | 1.60 | 1.34 – 1.90 | <0.001 |
| Genus: Klebsiella | 1.61 | 1.39 – 1.86 | <0.001 |
| Genus: Morganella | 0.81 | 0.58 – 1.13 | 0.207 |
| Used antibiotics | 1.26 | 1.11 – 1.44 | <0.001 |
| Had MRSA | 1.49 | 1.23 – 1.80 | <0.001 |
| Male | 1.32 | 1.18 – 1.47 | <0.001 |
| S(Culture drawn at ER) |  |  | 0.001 |
| Observations | 11592 |  |  |

**Table S5.** Estimates of *Male* model fitted to the original data. S(Male): The time-varying coefficient of *Male,* testing if this coefficient equals zero for all f; (/3): results presented per 3 unit increase in the covariate; (/5): results presented per 5 unit increase in the covariate.

| *Covariate* | **Male model** |  |  |
| --- | --- | --- | --- |
|  | *Odds Ratios* | *Conf. Int (95%)* | *P-Value* |
| Intercept | 0.09 | 0.07 – 0.12 | <0.001 |
| Age (/5) | 0.98 | 0.97 – 1.00 | 0.037 |
| Times hospitalized past year (/3) | 1.24 | 1.11 – 1.39 | <0.001 |
| Had Acinetobacter | 2.06 | 1.25 – 3.40 | 0.005 |
| Had Pseudomonas | 1.97 | 1.40 – 2.79 | <0.001 |
| Had CRE | 1.59 | 0.82 – 3.08 | 0.166 |
| Had VRE | 0.57 | 0.23 – 1.43 | 0.233 |
| Had ESBL | 2.62 | 2.32 – 2.96 | <0.001 |
| Had KPC | 1.81 | 1.24 – 2.64 | 0.002 |
| Culture drawn at the surgical unit | 0.94 | 0.80 – 1.10 | 0.466 |
| Culture drawn at the orthopedic unit | 1.38 | 1.11 – 1.72 | 0.004 |
| Culture drawn at the obstetrics unit | 1.11 | 0.60 – 2.05 | 0.731 |
| Culture drawn at other unit | 0.65 | 0.46 – 0.91 | 0.012 |
| Had catheter | 1.24 | 1.08 – 1.42 | 0.003 |
| Nosocomial | 1.27 | 1.12 – 1.44 | <0.001 |
| Arrived from an institution | 1.43 | 1.24 – 1.67 | <0.001 |
| Arrived from a medical clinic | 0.98 | 0.53 – 1.83 | 0.952 |
| Arrived from another hospital | 1.36 | 1.00 – 1.86 | 0.051 |
| Arrived from: unknown | 1.00 | 0.67 – 1.49 | 0.987 |
| Arrived from: other | 0.95 | 0.77 – 1.18 | 0.648 |
| Diabetes | 1.00 | 0.86 – 1.16 | 0.984 |
| Immunosuppression | 1.22 | 0.94 – 1.58 | 0.127 |
| Sample location: unknown | 0.64 | 0.53 – 0.78 | <0.001 |
| Sample location: sputum | 0.46 | 0.35 – 0.60 | <0.001 |
| Sample location: wound | 0.96 | 0.81 – 1.14 | 0.673 |
| Sample location: blood | 0.78 | 0.65 – 0.94 | 0.008 |
| Sample location: other | 0.70 | 0.27 – 1.81 | 0.460 |
| Genus: Entrobacter | 0.46 | 0.31 – 0.67 | <0.001 |
| Genus: Citrobacter | 0.10 | 0.04 – 0.24 | <0.001 |
| Genus: Pseudomonas | 0.81 | 0.69 – 0.96 | 0.013 |
| Genus: *Proteus* | 1.61 | 1.35 – 1.91 | <0.001 |
| Genus: *Klebsiella* | 1.61 | 1.39 – 1.86 | <0.001 |
| Genus: *Morganella* | 0.79 | 0.57 – 1.11 | 0.174 |
| Used antibiotics | 1.25 | 1.10 – 1.42 | <0.001 |
| Had MRSA | 1.51 | 1.25 – 1.82 | <0.001 |
| Culture drawn at ER | 1.38 | 0.89 – 2.13 | 0.151 |
| S(Male) |  |  | <0.001 |
| Observations | 11592 |  |  |

**Table S6.** Estimates of *Used antibiotics* model fitted to the original data. S(Used antibiotics): The time-varying coefficient of *Used antibiotics,* testing if this coefficient equals zero for all *t;* (/3): results presented per 3 unit increase in the covariate; (/5): results presented per 5 unit increase in the covariate.

| *Covariate* | **Used antibiotics model** |  |  |
| --- | --- | --- | --- |
|  | *Odds Ratios* | *Conf. Int (95%)* | *P-Value* |
| Intercept | 0.09 | 0.07 – 0.12 | <0.001 |
| Age (/5) | 0.98 | 0.97 – 1.00 | 0.026 |
| Times hospitalized past year (/3) | 1.25 | 1.12 – 1.39 | <0.001 |
| Had Acinetobacter | 2.02 | 1.22 – 3.33 | 0.006 |
| Had Pseudomonas | 1.94 | 1.37 – 2.73 | <0.001 |
| Had CRE | 1.51 | 0.79 – 2.90 | 0.214 |
| Had VRE | 0.64 | 0.26 – 1.57 | 0.325 |
| Had ESBL | 2.61 | 2.32 – 2.95 | <0.001 |
| Had KPC | 1.81 | 1.24 – 2.63 | 0.002 |
| Culture drawn at the surgical unit | 0.96 | 0.82 – 1.12 | 0.612 |
| Culture drawn at the orthopedic unit | 1.40 | 1.13 – 1.74 | 0.002 |
| Culture drawn at the obstetrics unit | 1.14 | 0.62 – 2.10 | 0.681 |
| Culture drawn at other unit | 0.65 | 0.47 – 0.91 | 0.012 |
| Had catheter | 1.26 | 1.10 – 1.45 | 0.001 |
| Nosocomial | 1.26 | 1.11 – 1.43 | <0.001 |
| Arrived from an institution | 1.44 | 1.24 – 1.67 | <0.001 |
| Arrived from a medical clinic | 0.95 | 0.51 – 1.78 | 0.882 |
| Arrived from another hospital | 1.34 | 0.98 – 1.82 | 0.065 |
| Arrived from: unknown | 0.98 | 0.66 – 1.47 | 0.928 |
| Arrived from: other | 0.98 | 0.79 – 1.21 | 0.849 |
| Diabetes | 1.02 | 0.88 – 1.18 | 0.767 |
| Immunosuppression | 1.23 | 0.95 – 1.59 | 0.119 |
| Sample location: unknown | 0.64 | 0.53 – 0.78 | <0.001 |
| Sample location: sputum | 0.45 | 0.35 – 0.59 | <0.001 |
| Sample location: wound | 0.97 | 0.82 – 1.14 | 0.692 |
| Sample location: blood | 0.77 | 0.65 – 0.93 | 0.006 |
| Sample location: other | 0.72 | 0.28 – 1.86 | 0.496 |
| Genus: Entrobacter | 0.46 | 0.31 – 0.67 | <0.001 |
| Genus: Citrobacter | 0.10 | 0.04 – 0.25 | <0.001 |
| Genus: Pseudomonas | 0.81 | 0.69 – 0.95 | 0.012 |
| Genus: Proteus | 1.61 | 1.35 – 1.91 | <0.001 |
| Genus: Klebsiella | 1.61 | 1.40 – 1.87 | <0.001 |
| Genus: Morganella | 0.80 | 0.58 – 1.12 | 0.191 |
| Had MRSA | 1.49 | 1.23 – 1.80 | <0.001 |
| Culture drawn at ER | 1.41 | 0.91 – 2.19 | 0.122 |
| Male | 1.32 | 1.18 – 1.48 | <0.001 |
| S(Used antibiotics) |  |  | 0.004 |
| Observations | 11592 |  |  |

**Table S7.** Estimates of *Had MRSA* model fitted to the original data. *S(Had MRSA)-.* The time-varying coefficient of *Had MRSA,* testing if this coefficient equals zero for all f; (/3): results presented per 3 unit increase in the covariate; (/5): results presented per 5 unit increase in the covariate

| *Covariate* | **Had MRSA model** |  |  |
| --- | --- | --- | --- |
|  | *Odds Ratios* | *Conf. Int (95%)* | *P-Value* |
| Intercept | 0.09 | 0.07 – 0.12 | <0.001 |
| Age (/5) | 0.98 | 0.97 – 1.00 | 0.027 |
| Times hospitalized past year (/3) | 1.24 | 1.11 – 1.38 | <0.001 |
| Had Acinetobacter | 2.05 | 1.24 – 3.39 | 0.005 |
| Had Pseudomonas | 1.96 | 1.39 – 2.77 | <0.001 |
| Had CRE | 1.56 | 0.82 – 3.00 | 0.177 |
| Had VRE | 0.62 | 0.25 – 1.56 | 0.312 |
| Had ESBL | 2.62 | 2.32 – 2.95 | <0.001 |
| Had KPC | 1.79 | 1.23 – 2.61 | 0.002 |
| Culture drawn at the surgical unit | 0.96 | 0.82 – 1.12 | 0.602 |
| Culture drawn at the orthopedic unit | 1.42 | 1.14 – 1.77 | 0.002 |
| Culture drawn at the obstetrics unit | 1.14 | 0.62 – 2.09 | 0.682 |
| Culture drawn at other unit | 0.65 | 0.47 – 0.91 | 0.012 |
| Had catheter | 1.25 | 1.09 – 1.44 | 0.001 |
| Nosocomial | 1.26 | 1.11 – 1.43 | <0.001 |
| Arrived from an institution | 1.43 | 1.23 – 1.66 | <0.001 |
| Arrived from a medical clinic | 0.96 | 0.52 – 1.79 | 0.910 |
| Arrived from another hospital | 1.33 | 0.98 – 1.81 | 0.070 |
| Arrived from: unknown | 0.98 | 0.66 – 1.47 | 0.939 |
| Arrived from: other | 0.97 | 0.78 – 1.20 | 0.787 |
| Diabetes | 1.03 | 0.89 – 1.19 | 0.696 |
| Immunosuppression | 1.23 | 0.95 – 1.59 | 0.112 |
| Sample location: unknown | 0.64 | 0.53 – 0.78 | <0.001 |
| Sample location: sputum | 0.45 | 0.35 – 0.59 | <0.001 |
| Sample location: wound | 0.96 | 0.81 – 1.14 | 0.633 |
| Sample location: blood | 0.78 | 0.65 – 0.93 | 0.006 |
| Sample location: other | 0.72 | 0.28 – 1.86 | 0.493 |
| Genus: Entrobacter | 0.45 | 0.31 – 0.67 | <0.001 |
| Genus: Citrobacter | 0.10 | 0.04 – 0.24 | <0.001 |
| Genus: Pseudomonas | 0.81 | 0.69 – 0.95 | 0.010 |
| Genus: Proteus | 1.61 | 1.35 – 1.91 | <0.001 |
| Genus: Klebsiella | 1.62 | 1.40 – 1.87 | <0.001 |
| Genus: Morganella | 0.81 | 0.58 – 1.13 | 0.208 |
| Used antibiotics | 1.26 | 1.11 – 1.43 | <0.001 |
| Culture drawn at ER | 1.41 | 0.91 – 2.19 | 0.120 |
| Male | 1.32 | 1.18 – 1.48 | <0.001 |
| S(Had MRSA) |  |  | <0.001 |
| Observations | 11592 |  |  |

**Table S8.** Estimates of *Culture drawn at ER* model with a random intercept fitted to the orginal data. S(Culture drawn at ER): The time-varying coefficient of *Culture drawn at ER,* testing if this coefficient equals zero for all f; (/3): results presented per 3 unit increase in the covariate; (/5): results presented per 5 unit increase in the covariate. The variance of the random intercept is 2.89 (95% confidence interval: 1.60, 1.90).

| *Covariate* | **Culture drawn at ER model** |  |  |
| --- | --- | --- | --- |
|  | *Odds Ratios* | *Conf. Int (95%)* | *P-Value* |
| Intercept | 0.05 | 0.03 – 0.07 | <0.001 |
| Age (/5) | 0.99 | 0.97 – 1.01 | 0.433 |
| Times hospitalized past year (/3) | 1.24 | 1.06 – 1.45 | 0.006 |
| Had Acinetobacter | 2.47 | 1.10 – 5.57 | 0.029 |
| Had Pseudomonas | 1.71 | 0.94 – 3.13 | 0.079 |
| Had CRE | 0.65 | 0.18 – 2.28 | 0.497 |
| Had VRE | 0.33 | 0.07 – 1.64 | 0.176 |
| Had ESBL | 3.40 | 2.83 – 4.08 | <0.001 |
| Had KPC | 1.64 | 0.86 – 3.12 | 0.130 |
| Culture drawn at the surgical unit | 1.05 | 0.84 – 1.31 | 0.658 |
| Culture drawn at the orthopedic unit | 1.41 | 1.03 – 1.93 | 0.031 |
| Culture drawn at the obstetrics unit | 1.10 | 0.49 – 2.48 | 0.820 |
| Culture drawn at other unit | 0.58 | 0.37 – 0.91 | 0.018 |
| Had catheter | 1.28 | 1.06 – 1.56 | 0.011 |
| Nosocomial | 1.24 | 1.05 – 1.46 | 0.009 |
| Arrived from an institution | 1.54 | 1.24 – 1.92 | <0.001 |
| Arrived from a medical clinic | 0.97 | 0.42 – 2.23 | 0.937 |
| Arrived from another hospital | 1.68 | 1.06 – 2.67 | 0.029 |
| Arrived from: unknown | 1.39 | 0.82 – 2.36 | 0.216 |
| Arrived from: other | 0.99 | 0.72 – 1.36 | 0.956 |
| Diabetes | 1.00 | 0.81 – 1.23 | 0.976 |
| Immunosuppression | 0.99 | 0.67 – 1.47 | 0.963 |
| Sample location: unknown | 0.59 | 0.46 – 0.76 | <0.001 |
| Sample location: sputum | 0.41 | 0.29 – 0.57 | <0.001 |
| Sample location: wound | 0.90 | 0.72 – 1.13 | 0.378 |
| Sample location: blood | 0.82 | 0.65 – 1.03 | 0.082 |
| Sample location: other | 0.82 | 0.27 – 2.52 | 0.733 |
| Genus: Entrobacter | 0.47 | 0.30 – 0.74 | 0.001 |
| Genus: Citrobacter | 0.08 | 0.03 – 0.22 | <0.001 |
| Genus: Pseudomonas | 0.80 | 0.65 – 0.98 | 0.030 |
| Genus: Proteus | 1.48 | 1.19 – 1.85 | 0.001 |
| Genus: Klebsiella | 1.76 | 1.46 – 2.13 | <0.001 |
| Genus: Morganella | 0.74 | 0.50 – 1.11 | 0.149 |
| Used antibiotics | 1.33 | 1.12 – 1.57 | 0.001 |
| Had MRSA | 1.52 | 1.13 – 2.07 | 0.007 |
| Male | 1.40 | 1.19 – 1.65 | <0.001 |
| S(Culture drawn at ER) |  |  | 0.026 |
| Random intercept |  |  | <0.001 |
| Observations | 11592 |  |  |

**Table S9.**
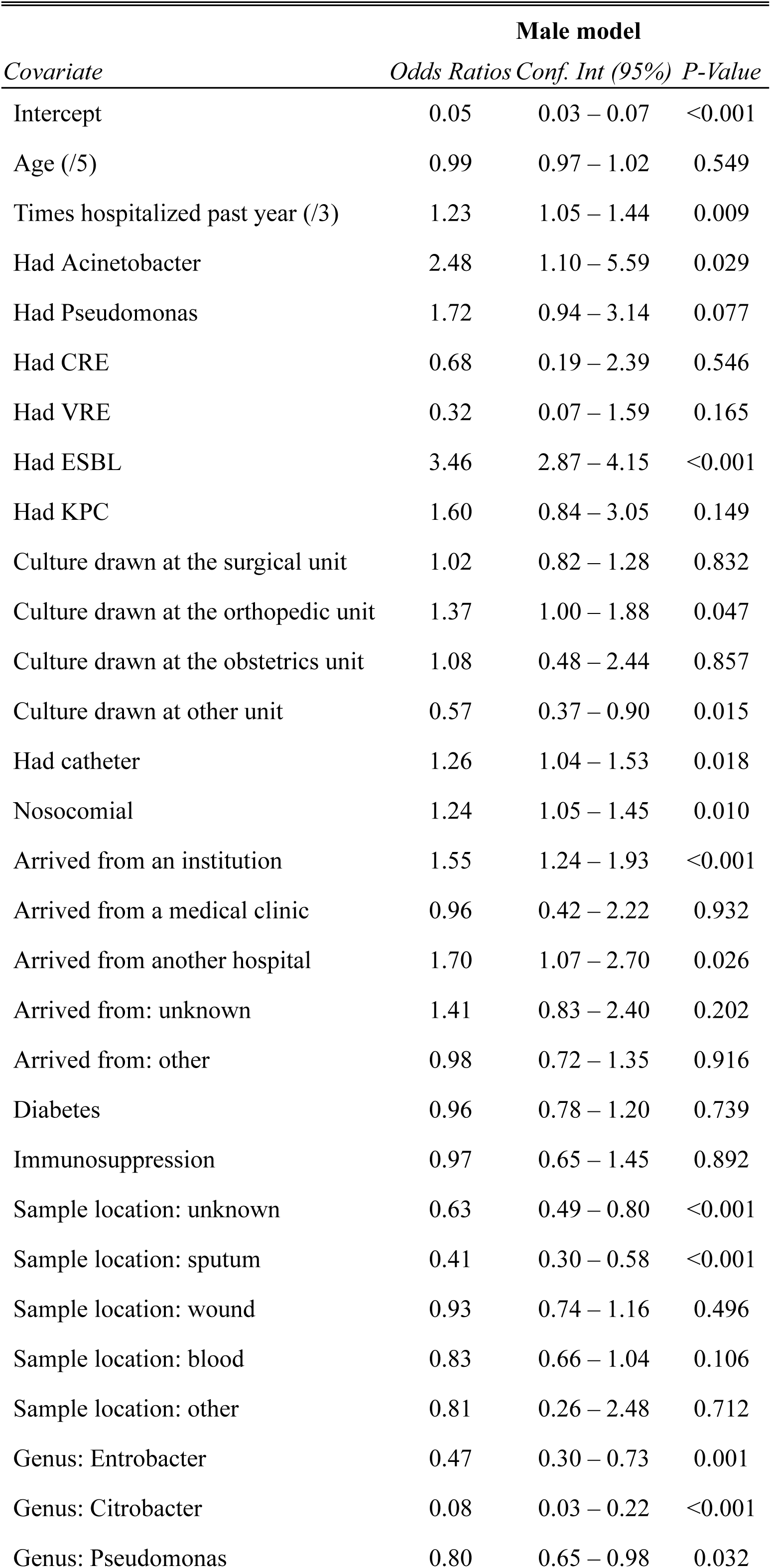
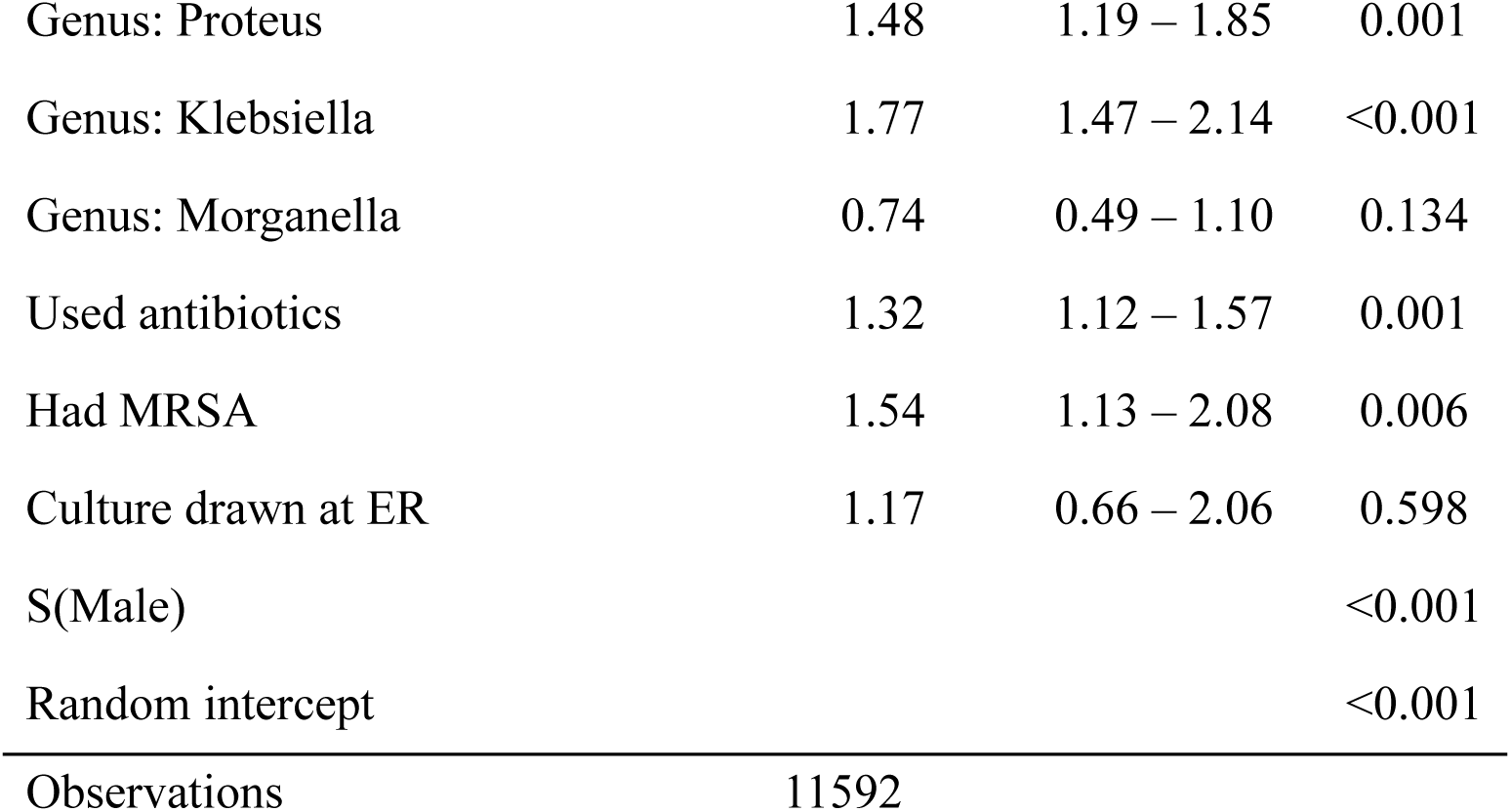
Estimates of *Male* model with a random intercept fitted to the orginal data. S(Male) The time-varying coefficient of *Male,* testing if this coefficient equals zero for all *t;* (/3): results presented per 3 unit increase in the covariate; (/5): results presented per 5 unit increase in the covariate. The variance of the random intercept is 2.89 (95% confidence interval: 1.60, 1.90).

**Table S10.** Estimates of *Used antibiotics* model with a random intercept fitted to the orginal data. S(Used antibiotics): The time-varying coefficient of *Used antibiotics,* testing if this coefficient equals zero for all f; (/3): results presented per 3 unit increase in the covariate; (/5): results presented per 5 unit increase in the covariate. The variance of the random intercept is 2.89 (95% confidence interval: 1.60, 1.90).

| *Covariate* | **Used antibiotics model** |  |  |
| --- | --- | --- | --- |
|  | *Odds Ratios* | *Conf. Int (95%)* | *P-Value* |
| Intercept | 0.05 | 0.03 – 0.07 | <0.001 |
| Age (/5) | 0.99 | 0.97 – 1.02 | 0.525 |
| Times hospitalized past year (/3) | 1.24 | 1.06 – 1.45 | 0.006 |
| Had Acinetobacter | 2.45 | 1.09 – 5.52 | 0.031 |
| Had Pseudomonas | 1.70 | 0.93 – 3.11 | 0.083 |
| Had CRE | 0.64 | 0.18 – 2.26 | 0.487 |
| Had VRE | 0.34 | 0.07 – 1.66 | 0.180 |
| Had ESBL | 3.41 | 2.84 – 4.10 | <0.001 |
| Had KPC | 1.64 | 0.86 – 3.11 | 0.132 |
| Culture drawn at the surgical unit | 1.04 | 0.84 – 1.30 | 0.708 |
| Culture drawn at the orthopedic unit | 1.40 | 1.02 – 1.91 | 0.036 |
| Culture drawn at the obstetrics unit | 1.11 | 0.49 – 2.52 | 0.798 |
| Culture drawn at other unit | 0.59 | 0.38 – 0.92 | 0.021 |
| Had catheter | 1.28 | 1.05 – 1.55 | 0.013 |
| Nosocomial | 1.23 | 1.05 – 1.45 | 0.011 |
| Arrived from an institution | 1.54 | 1.24 – 1.92 | <0.001 |
| Arrived from a medical clinic | 0.97 | 0.42 – 2.24 | 0.949 |
| Arrived from another hospital | 1.68 | 1.06 – 2.67 | 0.029 |
| Arrived from: unknown | 1.38 | 0.82 – 2.35 | 0.229 |
| Arrived from: other | 0.99 | 0.73 – 1.36 | 0.975 |
| Diabetes | 0.99 | 0.80 – 1.23 | 0.937 |
| Immunosuppression | 0.98 | 0.66 – 1.46 | 0.933 |
| Sample location: unknown | 0.62 | 0.49 – 0.80 | <0.001 |
| Sample location: sputum | 0.41 | 0.29 – 0.58 | <0.001 |
| Sample location: wound | 0.92 | 0.74 – 1.15 | 0.467 |
| Sample location: blood | 0.83 | 0.66 – 1.03 | 0.094 |
| Sample location: other | 0.82 | 0.27 – 2.53 | 0.736 |
| Genus: Entrobacter | 0.47 | 0.30 – 0.73 | 0.001 |
| Genus: Citrobacter | 0.08 | 0.03 – 0.22 | <0.001 |
| Genus: Pseudomonas | 0.80 | 0.65 – 0.98 | 0.034 |
| Genus: Proteus | 1.48 | 1.19 – 1.85 | <0.001 |
| Genus: Klebsiella | 1.77 | 1.47 – 2.13 | <0.001 |
| Genus: Morganella | 0.74 | 0.50 – 1.11 | 0.146 |
| Had MRSA | 1.52 | 1.12 – 2.06 | 0.007 |
| Culture drawn at ER | 1.20 | 0.68 – 2.12 | 0.539 |
| Male | 1.41 | 1.19 – 1.66 | <0.001 |
| S(Used antibiotics) |  |  | 0.006 |
| Random intercept |  |  | <0.001 |
| Observations | 11592 |  |  |

**Table S11.** Estimates of ***Had MRSA*** model with a random intercept fitted to the orginal data. S*(Had MRSA):* The time-varying coefficient of *Had MRSA.* testing if this coefficient equals zero for all f; (/3): results presented per 3 unit increase in the covariate; (/5): results presented per 5 unit increase in the covariate. The variance of the random intercept is 2.89 (95% confidence interval: 1.60, 1.90).

| *Covariate* | **Had MRSA model** |  |  |
| --- | --- | --- | --- |
|  | *Odds Ratios* | *Conf. Int (95%)* | *P-Value* |
| Intercept | 0.05 | 0.03 – 0.07 | <0.001 |
| Age (/5) | 0.99 | 0.97 – 1.02 | 0.533 |
| Times hospitalized past year (/3) | 1.24 | 1.06 – 1.44 | 0.007 |
| Had Acinetobacter | 2.47 | 1.09 – 5.58 | 0.030 |
| Had Pseudomonas | 1.73 | 0.95 – 3.16 | 0.075 |
| Had CRE | 0.65 | 0.18 – 2.32 | 0.512 |
| Had VRE | 0.30 | 0.06 – 1.55 | 0.150 |
| Had ESBL | 3.41 | 2.83 – 4.09 | <0.001 |
| Had KPC | 1.64 | 0.86 – 3.12 | 0.133 |
| Culture drawn at the surgical unit | 1.04 | 0.84 – 1.30 | 0.707 |
| Culture drawn at the orthopedic unit | 1.43 | 1.04 – 1.95 | 0.027 |
| Culture drawn at the obstetrics unit | 1.12 | 0.49 – 2.53 | 0.789 |
| Culture drawn at other unit | 0.59 | 0.38 – 0.93 | 0.022 |
| Had catheter | 1.27 | 1.05 – 1.54 | 0.014 |
| Nosocomial | 1.23 | 1.04 – 1.44 | 0.013 |
| Arrived from an institution | 1.55 | 1.24 – 1.92 | <0.001 |
| Arrived from a medical clinic | 0.99 | 0.43 – 2.27 | 0.978 |
| Arrived from another hospital | 1.68 | 1.05 – 2.67 | 0.029 |
| Arrived from: unknown | 1.38 | 0.81 – 2.34 | 0.232 |
| Arrived from: other | 1.00 | 0.73 – 1.37 | 0.982 |
| Diabetes | 1.00 | 0.81 – 1.24 | 0.990 |
| Immunosuppression | 0.98 | 0.66 – 1.46 | 0.932 |
| Sample location: unknown | 0.62 | 0.49 – 0.79 | <0.001 |
| Sample location: sputum | 0.41 | 0.30 – 0.58 | <0.001 |
| Sample location: wound | 0.92 | 0.73 – 1.15 | 0.457 |
| Sample location: blood | 0.83 | 0.66 – 1.03 | 0.093 |
| Sample location: other | 0.82 | 0.27 – 2.53 | 0.736 |
| Genus: Entrobacter | 0.47 | 0.30 – 0.73 | 0.001 |
| Genus: Citrobacter | 0.08 | 0.03 – 0.22 | <0.001 |
| Genus: Pseudomonas | 0.80 | 0.65 – 0.98 | 0.032 |
| Genus: Proteus | 1.49 | 1.19 – 1.85 | <0.001 |
| Genus: Klebsiella | 1.78 | 1.47 – 2.14 | <0.001 |
| Genus: Morganella | 0.75 | 0.50 – 1.12 | 0.161 |
| Used antibiotics | 1.33 | 1.12 – 1.57 | 0.001 |
| Culture drawn at ER | 1.20 | 0.68 – 2.13 | 0.526 |
| Male | 1.40 | 1.19 – 1.66 | <0.001 |
| S(Had MRSA) |  |  | 0.026 |
| Random intercept |  |  | <0.001 |
| Observations | 11592 |  |  |

**Figure S1.**
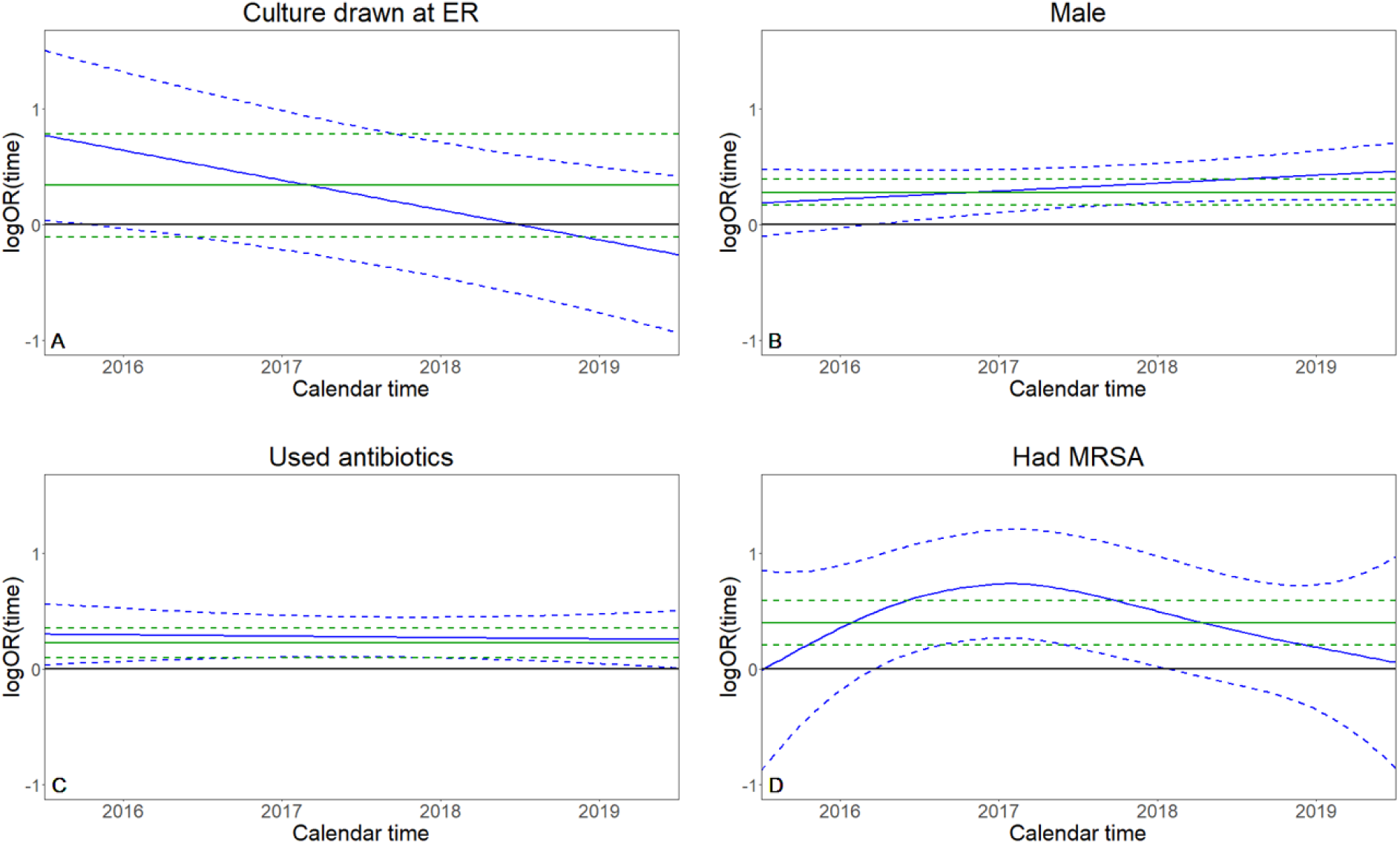
Estimates of the time-fixed (green) and time-varying (blue) coefficients from the four time-varying coefficient models with random intercepts and the single time-fixed model. Correspondingly, 95% Bayesian credible intervals and standard 95% confidence intervals are given in dashed lines. The horizontal axis represents the time between 2016 and 2019.

**Figure S2.**
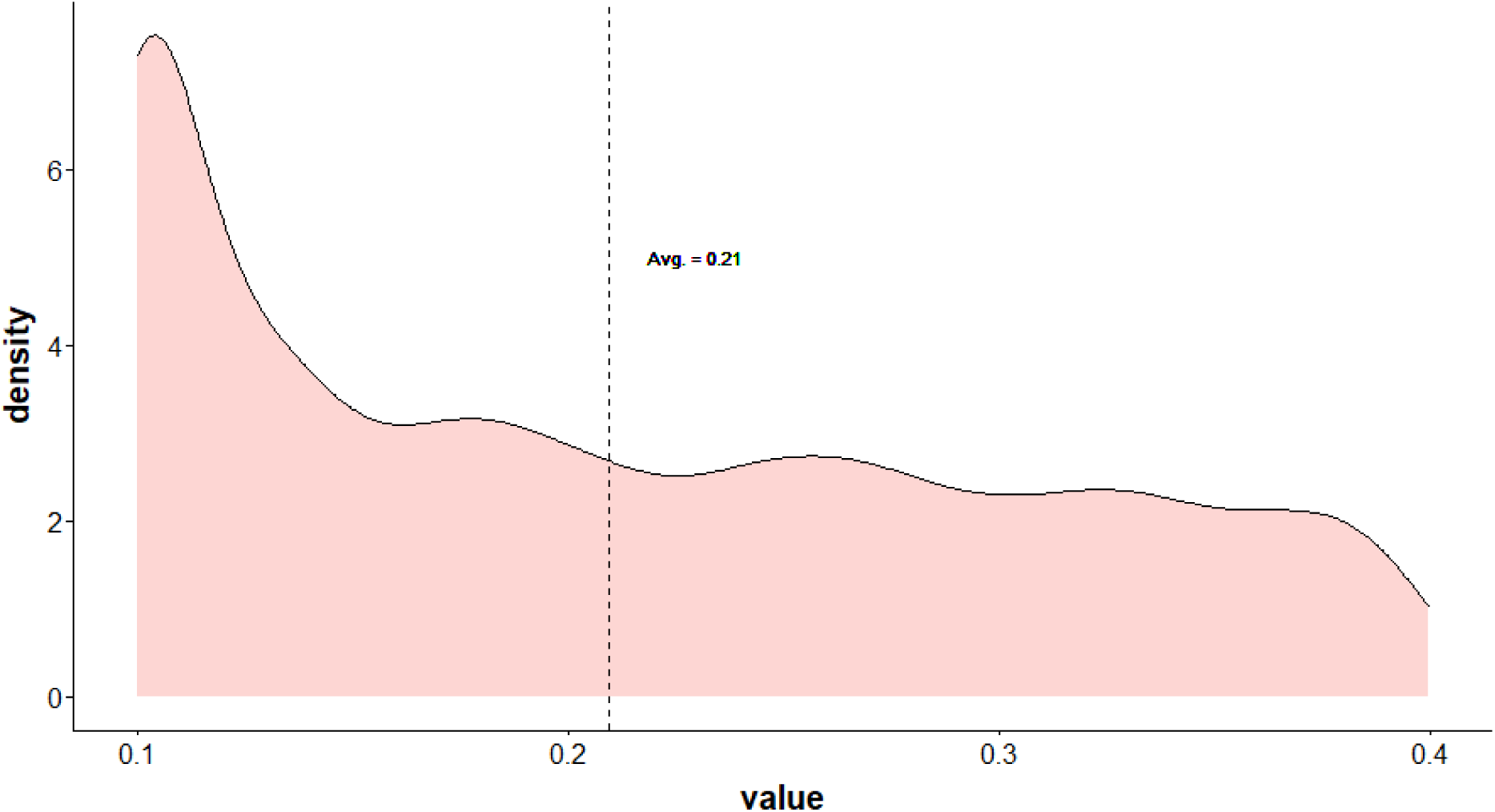
Density of *p_t_*(*Community use* = 1) in our data, first simulation study. The values of *t* are the withdrawal times of bacterial cultures from the patients.

**Figure S3.**
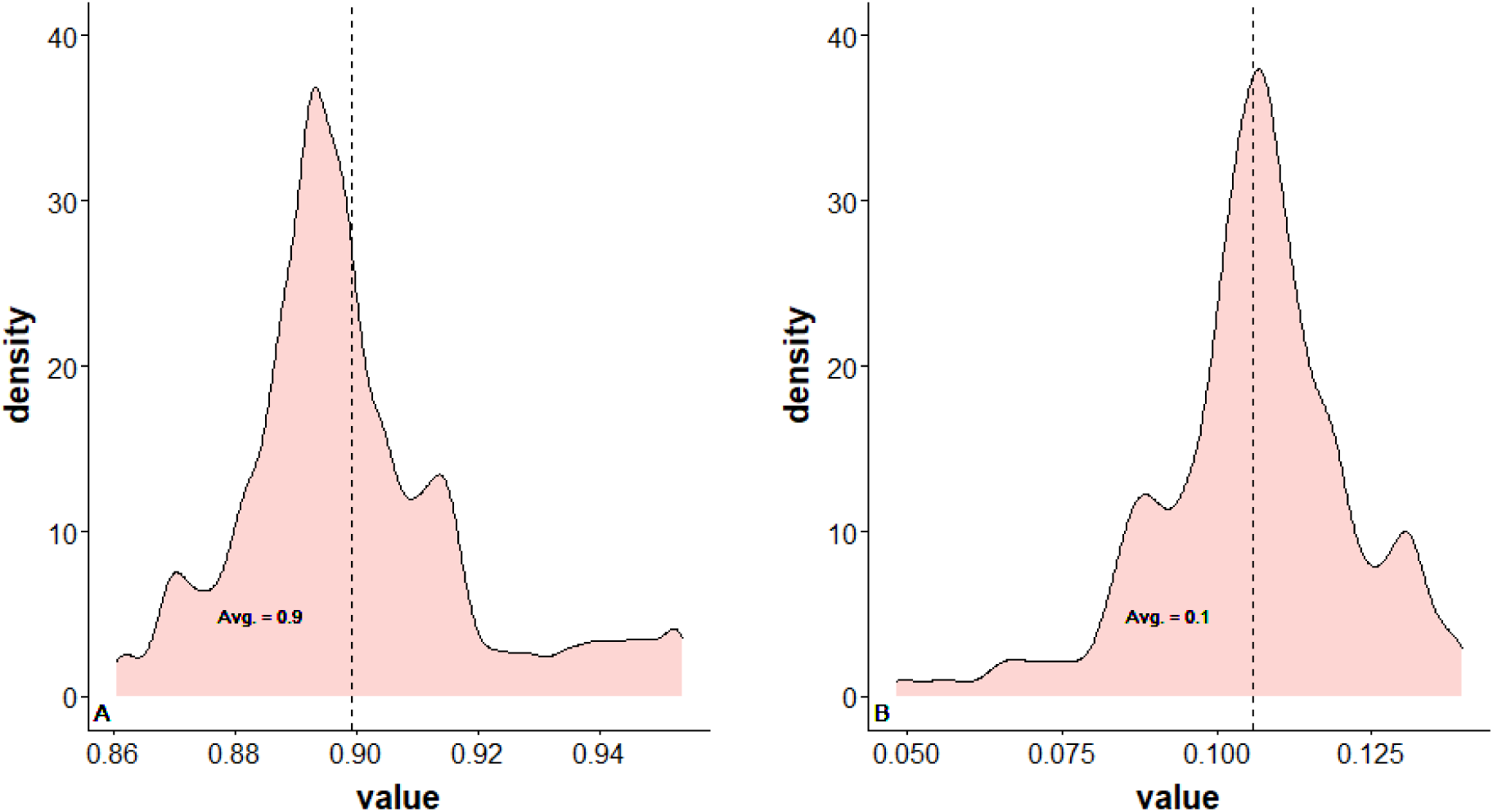
Density of *p_t_*(*Arrived at ER* = *k*) in our data, where *k* ∈ {0,1}, first simulation study. Panel (A): The estimated density of *p_t_*(*Arrived at ER* = 0). Panel (B): The estimated density of *p_t_*(*Arrived at ER* = 1). The values of *t* are the withdrawal times of bacterial cultures from the patients. The probability *p_t_*(*Arrived at ER* = *k*) was estimated from the Meir hospital data using a GAM.

**Figure S4.**
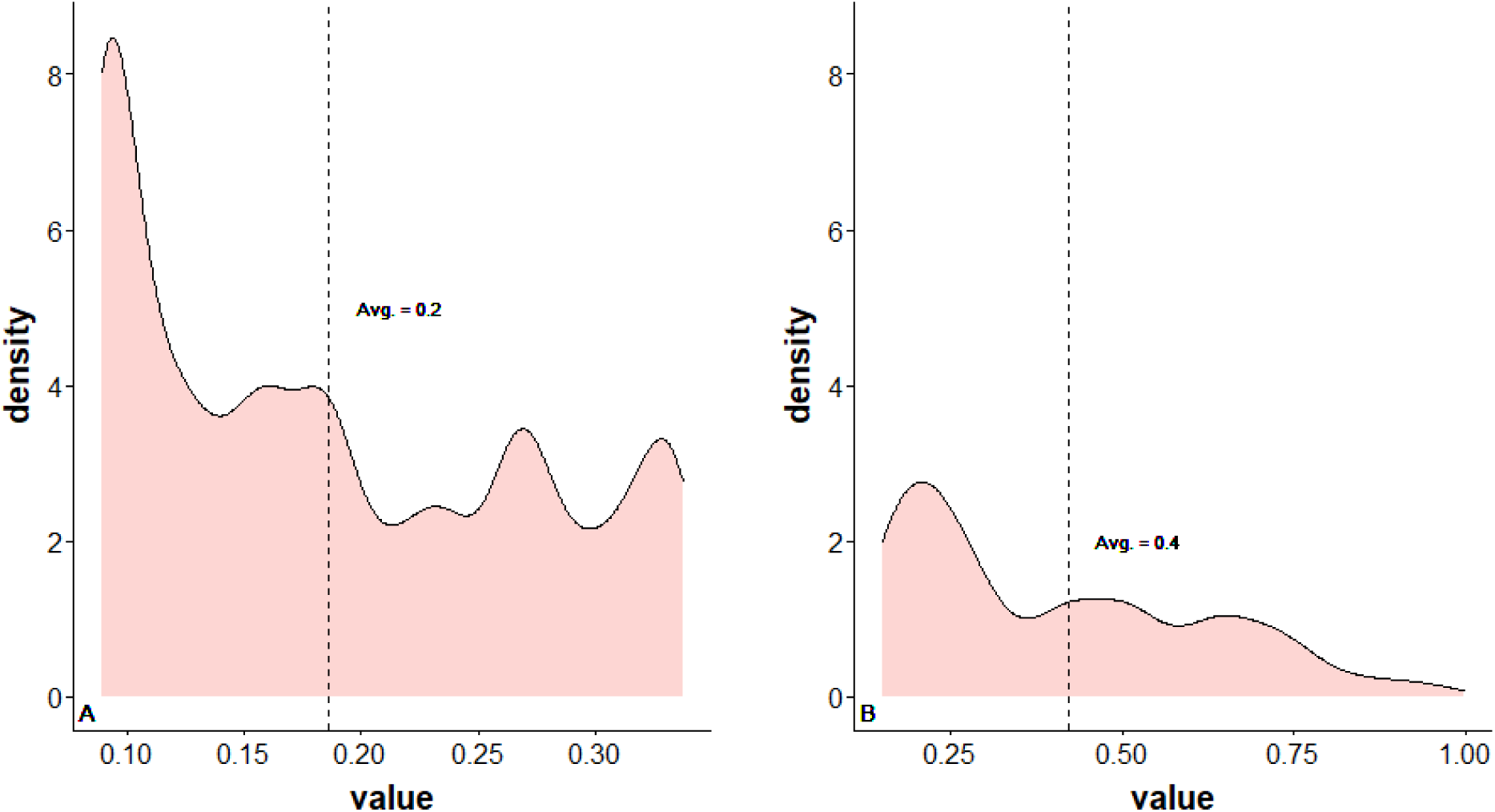
Density of *p_t_*(*Community use* = 1|*Arrived at ER* = *k*) in our data, where *k* ∈ {0,1} from Equation (3), first simulation study. Panel (A): The estimated density of *p_t_*(*Community use* = 1|*Arrived at ER* = 0). Panel (B): The estimated density of *p_t_*(*Community use* = 1|*Arrived at ER* = 1). The values of *t* are the withdrawal times of bacterial cultures from the patients. In the 15 rare instances when the sampled *p_t_*(*Community use* = 1|*Arrived at ER* = *k*) > 1, they were replaced by a uniform sample from [0.85, 1].

